# Genomic assessment of invasion dynamics of SARS-CoV-2 Omicron BA.1

**DOI:** 10.1101/2023.01.02.23284109

**Authors:** Joseph L.-H. Tsui, Ben Lambert, Sumali Bajaj, John T. McCrone, Rhys P.D. Inward, Paolo Bosetti, Verity Hill, Rosario Evans Pena, Alexander E. Zarebski, Thomas P. Peacock, Luyang Liu, Neo Wu, Megan Davis, Isaac I. Bogoch, Kamran Khan, Rachel Colquhoun, Áine O’Toole, Ben Jackson, Abhishek Dasgupta, Eduan Wilkinson, Houriiyah Tegally, Tulio de Oliveira, The COVID-19 Genomics UK (COG-UK) consortium, Thomas R. Connor, Nicholas J. Loman, Vittoria Colizza, Christophe Fraser, Erik Volz, Xiang Ji, Marc A. Suchard, Bernardo Gutierrez, Meera Chand, Simon Dellicour, Simon Cauchemez, Jayna Raghwani, Philippe Lemey, Andrew Rambaut, Oliver G. Pybus, Moritz U.G. Kraemer

## Abstract

SARS-CoV-2 variants of concern (VOCs) arise against the backdrop of increasingly heterogeneous human connectivity and population immunity. Through a large-scale phylodynamic analysis of 115,622 Omicron genomes, we identified >6,000 independent introductions of the antigenically distinct virus into England and reconstructed the dispersal history of resulting local transmission. Travel restrictions on southern Africa did not reduce BA.1 importation intensity as secondary hubs became major exporters. We explored potential drivers of BA.1 spread across England and discovered an early period during which viral lineage movements mainly occurred between larger cities, followed by a multi-focal spatial expansion shaped by shorter distance mobility patterns. We also found evidence that disease incidence impacted human commuting behaviours around major travel hubs. Our results offer a detailed characterisation of processes that drive the invasion of an emerging VOC across multiple spatial scales and provide unique insights on the interplay between disease spread and human mobility.

**Highlights:**

- Over 6,000 introductions ignited the epidemic wave of Omicron BA.1 in England
- Importations prior to international travel restrictions were responsible for majority of local BA.1 infections but importations continued from sources other than southern Africa
- Human mobility at regional and local spatial scales shaped dissemination and growth of BA.1
- Changes in human commuting patterns are associated with higher case incidence in travel hubs across England

## Introduction

Since the emergence of SARS-CoV-2 in late 2019, multiple variants of concern (VOCs) have sequentially dominated the pandemic across different regions in the world. The Omicron variant (PANGO lineage B.1.1.529 later divided into lineages including BA.1 and BA.2) was discovered first through genomic surveillance in Botswana and South Africa in late November 2021 (*1*) and designated a VOC by the World Health Organisation on 26 November 2021 (*2*). A dramatic initial surge in Omicron cases in South Africa suggested a higher transmission rate than previous variants (*3*), which studies later attributed to a shorter serial interval, increased immune evasion and greater intrinsic transmissibility than Delta (*4*–*7*). The mechanism for this greater transmissibility is hypothesised to be altered tropism and higher replication in the upper respiratory tract (*8, 9*). Together with a waning level of population immunity from previous infections and vaccination (*10*), local transmission of Omicron BA.1 was soon reported thereafter in major travel hubs worldwide, including New York City and London by early December 2021, despite travel restrictions on international flights from multiple southern African countries (*11, 12*).

Following the first confirmed case of Omicron BA.1 in England on 27 November 2021 (*13*), Omicron prevalence increased rapidly across all English regions, with Greater London peaking first in mid-December at ∼6% prevalence, followed by the South East region (*14*). Other metropolitan areas in the North West and North East, e.g. Greater Manchester and Newcastle, saw similar but delayed increases in prevalence with observed peaks between early- and mid-January 2022. Incidence of Omicron BA.1 had substantially declined in Greater London and other southern regions by early-January 2022, resulting in a gradient of decreasing prevalence from north to south England (*15*).

This spatiotemporal pattern of early BA.1 spread has some similarity with that of the Alpha variant (*16*), but is markedly different from the invasion of the Delta variant which spread initially from the North West and surrounding regions of England (*17*). The rapid surge in the number of infections during the initial emergence of Omicron in England prompted the UK government to impose a number of interventions, including a move to “Plan B” non-pharmaceutical restrictions (e.g. mandatory COVID pass for entry into certain indoor venues, face coverings and work-from-home guidance) on 8 December 2021 (*18*) and an accelerated vaccine rollout to provide access to booster doses for all adults by mid-December 2021 (*19*). From mid- to late-January 2022, the prevalence of SARS-CoV-2 in England decreased, coincident with a falling proportion of BA.1 infections, and the BA.2 lineage started to replace BA.1 as the dominant variant which was later replaced by BA.4 and BA.5 (*20*–*22*).

Understanding and quantifying the relative contributions of the various factors that shaped the spatial dissemination of Omicron BA.1 in England can help inform the design of spatially targeted interventions against future VOCs (*23*). Here we analyse the Omicron BA.1 wave in England, from its first detection in November 2021 to the end of January 2022. Specifically, we use a dataset of 48,615 Omicron BA.1 genomes sampled in England, corresponding to ∼1 percent of all confirmed COVID-19 cases of the Omicron variant in England during the study period and representing 313 lower tier local authorities (LTLAs), together with sub-city level aggregated and anonymized human mobility and epidemiological data. Furthermore, using a statistical model we explore the impact of changes in disease prevalence on the connectivity to and from major travel hubs in England.

## Results

### International seeding events and Omicron BA.1 lineage dynamics

To evaluate the timing of virus importations into England and the dynamics of their descendent local transmission lineages, we undertook a large-scale phylodynamic analysis of 115,622 SARS-CoV-2 Omicron genomes (BA.1/BA.2 and their descendent lineages) sampled globally between 8 November 2021 and 31 January 2022. About 42% (n=48,615) of the genomes were sampled from England and sequenced by the COVID-19 Genomics UK (COG-UK) consortium (*24*). All English and global genomes (as available from COG-UK and GISAID on 12 and 9 April 2022 respectively) collected prior to 28 November 2021 were included; genomes collected after that date were randomly subsampled in proportion to weekly Omicron case incidence while maintaining a roughly 1:1 ratio between England and non-England samples. Furthermore, to reduce sampling bias due to heterogeneous sequencing coverage, we performed a weighted subsampling of the English genomes using a previously developed procedure which takes into account differences in number of sequences sampled per reported case at the Upper Tier Local Authority (UTLA) level (https://github.com/robj411/sequencing_coverage) (Materials and Methods).

We identified at least 6,455 [95% HPD: 6,184 to 6,722] independent introduction events. Most importations from outside of England (69.9% [95% HPD: 69.0 to 70.7]) were singletons (i.e. a single English genome associated with an importation event that did not lead to any local transmission observable in our dataset). The earliest introduction event is estimated between 5 November and 18 November (approximated as the midpoint between the inferred times of the most recent common ancestor (MRCA) of the transmission lineage and the parent of the MRCA (TPMRCA)). Between the first introduction event and mid-December 2021, we infer an approximately exponential increase in the daily number of imports before a plateau in early January 2022 (Figure 1C). This rapid increase in importations occurred despite restrictions on incoming international travel from 11 southern African countries and was likely due in part to a surge of Omicron infections in US East Coast cities in early December 2021 and substantially greater influx of air passengers from the US. To derive city-level estimates of importation intensity from the US and country-level estimates from South Africa, we calculate the Estimated Importation Intensity (EII) using a combination of the weekly number of reported COVID-19 cases, weekly prevalence of Omicron BA.1 genomes, and monthly number of air passengers travelling to the United Kingdom from the origin airports (see Methods & Materials). Whilst the majority of early imports were likely to have come from South Africa, we find the average weekly Omicron BA.1 EII from the US was about four times that of South Africa during the weeks of travel restrictions; Figure 2).

**Figure 1:**
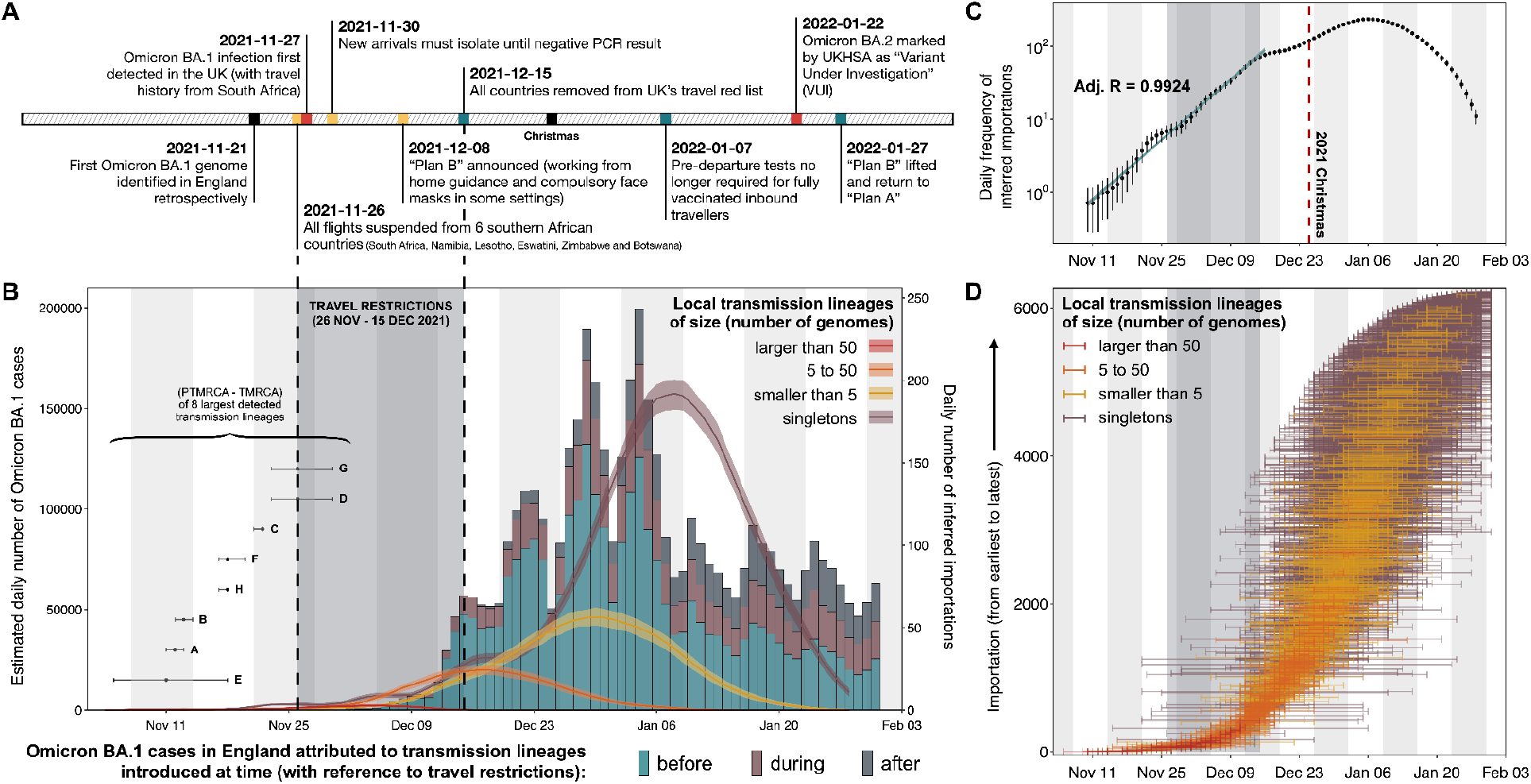
Dynamics of Omicron BA.1 transmission lineages in England. (A) Timeline of key events during the SARS-CoV-2 Omicron BA.1 wave in England until February 2022. (B) Histogram of the estimated daily number of Omicron BA.1 cases, coloured according to the proportion of cases that can be attributed to importations before/during/after the travel restrictions between 26 November and 15 December 2021 (shaded region). Solid lines represent the daily frequency of inferred importations (7-day rolling average), coloured according to the size of resulting local transmission lineages; shading denotes the 95% HPD across the posterior tree distribution. Forest plot (bottom left) shows the distribution of inferred time of importation, TPMRCA (inferred time of parent of most recent common ancestor) and TMRCA (inferred time of most recent common ancestor) of the 8 largest detected transmission lineages (labelled A to H). (C) Daily frequency of inferred importations (7-day rolling average), without stratification by size of resulting local transmission lineage (black dots); error bars denote the 95% HPD across the posterior tree distribution. Solid blue line represents the daily number of importations expected from an exponential model fitted to the observed 7-day rolling average importation intensity (Materials and Methods). (D) Distribution of TPMRCAs and TMRCAs of all 6,455 detected introductions. Each horizontal line represents a single introduction event that led to either a transmission lineage or a singleton, with the left limit indicating the TPMRCA and the right limit indicating the TMRCA (or the collection date of the genome sample for a singleton).

**Figure 2:**
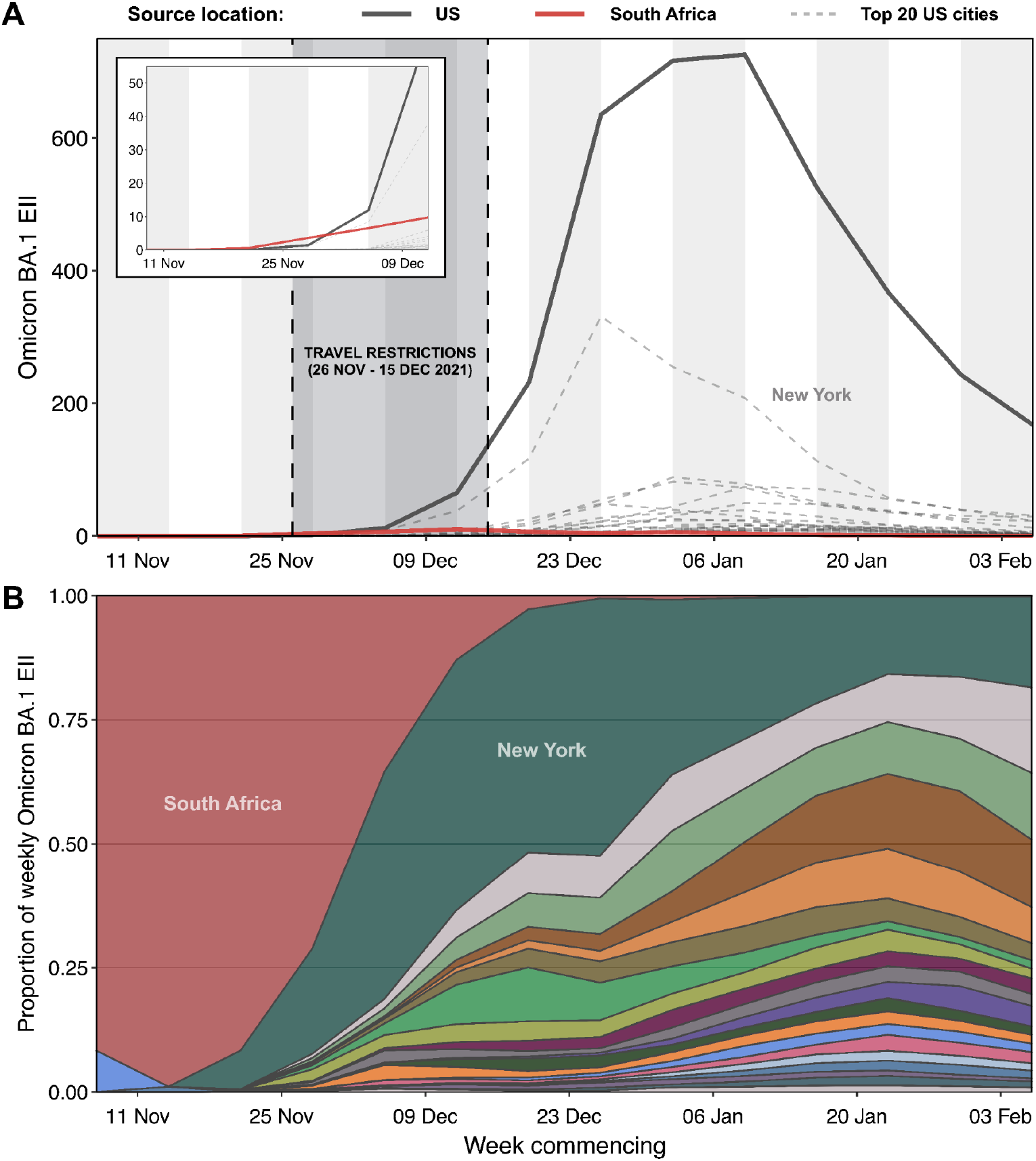
Importation intensity of Omicron BA.1 from South Africa and the United States. (A) Estimated weekly number of Omicron BA.1 cases arriving in the United Kingdom (Estimated Importation Intensity (EII); see Methods & Materials for calculations) from South Africa (red solid line) and 20 cities in the United States (each grey dashed line represents a city; black solid line represents the total EII from the 20 cities considered). Grey shaded region represents the period (26 November to 15 December 2021) when travel restrictions on international arrivals from multiple southern African countries were implemented. (B) Proportion of weekly EII of Omicron BA.1 by country and 20 cities in the United States.

To cross-validate the importation dynamics inferred from viral genomes using an independent data source, we extracted data from the Variant and Mutations (VAM) line list (*26*) provided by the UK Health Security Agency (UKHSA) to calculate the daily number of incoming travellers who later tested positive for Omicron in community surveillance (Pillar 2) of the UK SARS-CoV-2 testing programme. The temporal profile of importation intensity from these epidemiological data appears broadly consistent with that inferred from the phylodynamic analysis, with the latter being expanded and lagging in time compared to the former (Figure S1). This observation is consistent with previous studies (*27*), where the apparent discrepancy represents an increasing time lag between lineage importation into England and the time of first detection of local transmission events. This increase in importation lag is also evident from the increasing difference between the TPMRCA and TMRCA of transmission lineages as the epidemic progressed (Figure 1D).

As with the introduction and spread of previous variants in England (*17, 27*), we find overdispersion in transmission lineage sizes (Figure S2), with most sampled genomes belonging to a small number of large transmission lineages: the 8 largest transmission lineages identified (each comprising >700 genomes) combined accounts for >60% of English genomes in our dataset (Figure 1B). Six out of the 8 largest transmission lineages are inferred to have been imported before the travel restrictions were introduced (Figure 1B). Additionally, we observe a strong size-dependency on the time of importation of transmission lineages, with most large transmission lineages attributed to early seeding events detected between 5 and 13 November 2021 (Figure 1A). This characteristic can be recapitulated using a simple mathematical model under the assumption that all lineages share the same transmission characteristics except the timing of introduction (Materials and Methods), suggesting that timing of introduction is indeed the main determinant of transmission lineage size.

We infer that 399 transmission lineages (including the aforementioned 8 largest lineages) resulted from importations that occurred before the end of the travel restrictions on incoming flights from southern Africa (15 December 2021); 29 of these lineages were introduced before the travel restrictions were implemented (from 26 November 2021). Although these early importations account for only a small proportion (∼6%) of the estimated total number of introductions, they are responsible collectively for ∼80% of Omicron BA.1 infections reported in England, up to the end of January 2022. Further, early travel restrictions targeted at southern African countries were largely ineffective at reducing the overall intensity of importations,with continued exponential growth of importations through mid-December, partly due to a shift in source of importations of BA.1 away from South Africa to the US as shown using air passenger data (Figure 1B; Figure 2). Whilst the local epidemic may have been delayed had the travel restrictions been implemented earlier, importations from secondary hubs with unrestricted air traffic would have likely been responsible for the majority of local Omicron BA.1 infections.

Some transmission lineages from early importations were only detected several weeks after their inferred time of importation. However, caution is required when interpreting this result, as we cannot exclude the possibility that these transmission clusters represent the aggregation of multiple independent transmission lineages as a result of unsampled genetic diversity outside of England. Such aggregation of multiple transmission lineages would have resulted in earlier estimated dates of importation, potentially explaining the smaller than expected size (compared to predictions from simulations; Figure S3) of these transmission lineages with unusually long importation lag (*27*). Analyses incorporating detailed metadata on travel history could explore this question further (*28, 29*).

### Human mobility drives spatial expansion and heterogeneities in Omicron BA.1 growth

The rapid increase in Omicron importations between the beginning of November and mid-December 2021 led to the establishment of local transmission chains, initially concentrated in Greater London and neighbouring LTLAs in the South West region and East of England. This coincided with early increases in Omicron BA.1 prevalence in the corresponding regions, as observed from S-gene target failure (SGTF, a proxy for Omicron BA.1) data and other epidemiological studies (*15*). To further investigate the spatiotemporal dynamics of Omicron transmission lineages in England, we reconstructed the dispersal history of all identified transmission lineages (with >5 genomes) using spatially explicit phylogeographic techniques. Sampling of English genomes is highly representative of the estimated number of Omicron BA.1 cases at the UTLA level (Figure S4), therefore allowing unbiased reconstruction of the viral lineage movements across space.

We observe multiple distinct stages to the spread of BA.1 across England, with the 8 largest identified transmission lineages sharing broadly similar patterns of spatial dispersal. Unlike with other variants, we find a fairly even regional spread in the numbers of transmission lineages first detected in each region, with only ∼20% in Greater London (followed by 15.4% in South East and 13.3% in North West; and if only introductions prior to December 2021 are considered, the value for Greater London is 27.3%). However, most of the early cases outside of Greater London resulted in only limited local diffusion; widespread transmission developed concurrently with the rapid expansion centred in Greater London and surrounding areas by the end of November 2021 (Figure 3; Figure S5 and S6).

**Figure 3:**
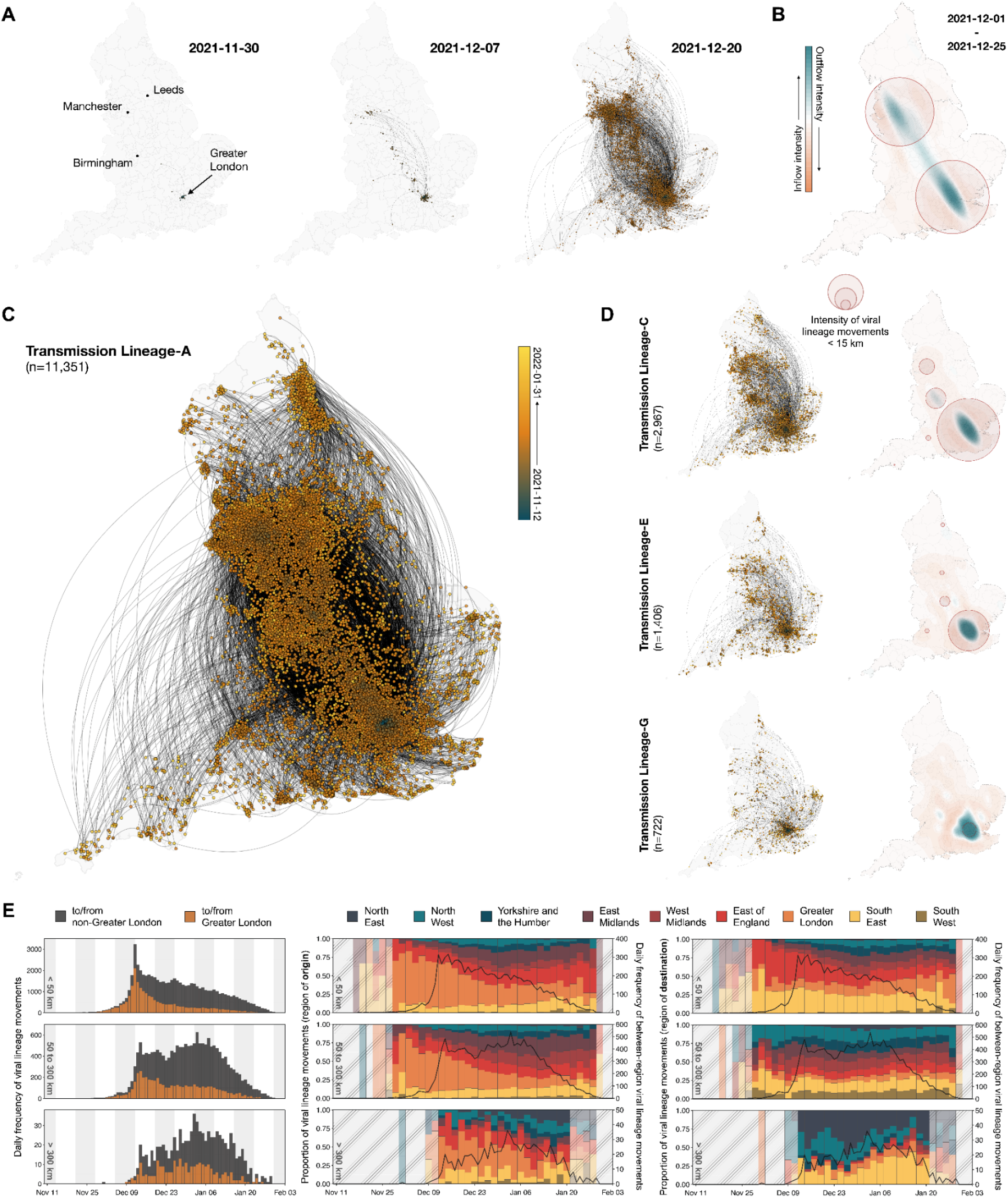
Spatiotemporal dynamics of Omicron BA.1 transmission lineages in England. (A and C) Continuous phylogeographic reconstruction of the dispersal history of the largest detected BA.1 transmission lineages in England (Transmission Lineage-A) across multiple time-points. Nodes are coloured according to inferred date of occurrence and the direction of viral lineage movement is indicated by edge curvature (anti-clockwise). (B) Geographical distribution of intensity of inflow and outflow of BA.1 viral lineages for transmission lineage Transmission Lineage-A. Blue colours indicate areas with high intensity of domestic exportation of viral lineages; red colours, high intensity of importation. Red circles indicate areas with high densities of local viral movements over distances <15 km, with larger radii representing higher densities. (D) Continuous phylogeographic reconstruction of Transmission Lineage-C, E, and G with corresponding maps showing the geographical distribution of intensity of domestic lineage movements (Transmission Lineages-B, D, F and H are shown in Figure S6). (E) Plots in each row correspond to viral lineage movements across different spatial scales (top: <50 km, middle: 50 to 300 km, bottom: >300 km). (Left) The histograms show the daily frequency of viral lineage movements across the different spatial scales. Colours indicate whether the origin and/or destination of the viral lineage movements are inferred to have occurred in Greater London. (Middle/Right) Solid black lines represent the daily frequency of between-region viral lineage movements across the different spatial scales. Vertical bars indicate the proportions of viral lineage movements (aggregated at 2-day intervals) with origin/destination in different regions as indicated by their corresponding colours. Shaded grey areas indicate periods when there were fewer than 9 inferred viral lineage movements per day at the corresponding spatial scale.

The frequency of long-distance (>300 km) viral lineage movements increased substantially during the first week of December, predominantly from Greater London to LTLAs across East/West Midlands and North West England. This pattern of strongly directional movement is most notable in the two largest transmission lineages (Transmission Lineage-A and Transmission Lineage-B), in which BA.1 genomes were first sampled in multiple urban conurbations including Nottingham, Leeds and Birmingham in early/mid-December 2021 (Figure 3A). However, local transmission was not established immediately in these cities and the ratio of local (within-city) versus all viral movements remained between 25% and 50% from December 2021 to January 2022, before climbing again when local mobility levels recovered after the holidays (*30*–*33*). In contrast, local viral movements in Greater London and Greater Manchester comprised about 90% and 60% of all movements between November and December 2021, respectively (Figure S5). The intensity of viral lineage movements from Greater London (with contributions from South East and East of England) continued to increase before plateauing in mid-December. This observation coincided with the early peak in Omicron BA.1 cases in some LTLAs in Greater London and other southern England regions discussed later in this manuscript (Figure 5A). Further, we observe a limited number of bidirectional long-distance viral movements between North West England and Greater London, with North West England acting primarily as a sink throughout the BA.1 wave (e.g. Transmission Lineage-A and Transmission Lineage-B); similar dynamics are also seen for the South West (Figure 3E).

We inferred a gradual but consistent decline through time in the frequency of short-range (<50 km) viral lineage movements, with Greater London acting as the predominant source of (cross-region) local diffusion for most of the epidemic (Figure 3E). Interestingly, we identify a second phase during which there was a notable increase in the frequency of mid-to-long range (50 to 300 km) viral movements (during the 2021/22 New Year period; Figure 3E, left). This period is also characterised by more frequent outward viral lineage movement from East Midlands, West Midlands and East of England, and more frequent inward movements in North East and North West England, whilst the relative contribution of Greater London to the onward propagation of BA.1 across England continued to diminish (Figure 3E, middle and right). This observation is consistent with epidemiological data, which show most areas outside of southern England experiencing a peak in Omicron BA.1 case incidence only by the first week of January 2022 (Figure 5B and C).

To assess and quantify the contribution of various demographic, epidemiological and mobility-related factors in shaping the dissemination of Omicron BA.1 in England, we use a discrete phylogeographic generalised linear model (GLM) approach to test their association with viral lineage movements among LTLAs (Materials and Methods). We fit this high-dimensional phylogeographic model to the three largest transmission lineages independently and jointly to the next five largest transmission lineages together comprising 61.9% of all English genomes (Materials and Methods). We allow the predictor support and effect size to vary temporally between the expansion phase (before 26 December 2021) and the post-expansion phase (26 December 2021 up to the end of January 2022) (*31, 32, 34*). This time-inhomogeneous model provides evidence for a dynamic spatial transmission process through time, with substantial changes in the effect size estimate for most of the predictors across the two phases (Figure 4B). We observe consistently strong support in the expansion phase for the gravity model - a spatial interaction model in which the intensity of travel between regions increases with population size at the origin and destination but decreases with distance between them, likely shaping early viral transmission in and around population centres. Consistent with results from continuous phylogeography, the early expansion is also characterised by strongly directional viral dissemination as indicated by the predictor support for a Greater London LTLA as origin as well as higher dissemination out of LTLAs with earlier peak times in three out of four analyses (Figure 4B; Figure S7). The effect size for a Greater London origin is particularly pronounced for smaller transmission lineages, again consistent with continuous phylogeography which shows smaller transmission lineages to be more localised near Greater London compared to their larger counterparts (Figure 3; Figure S6).

**Figure 4:**
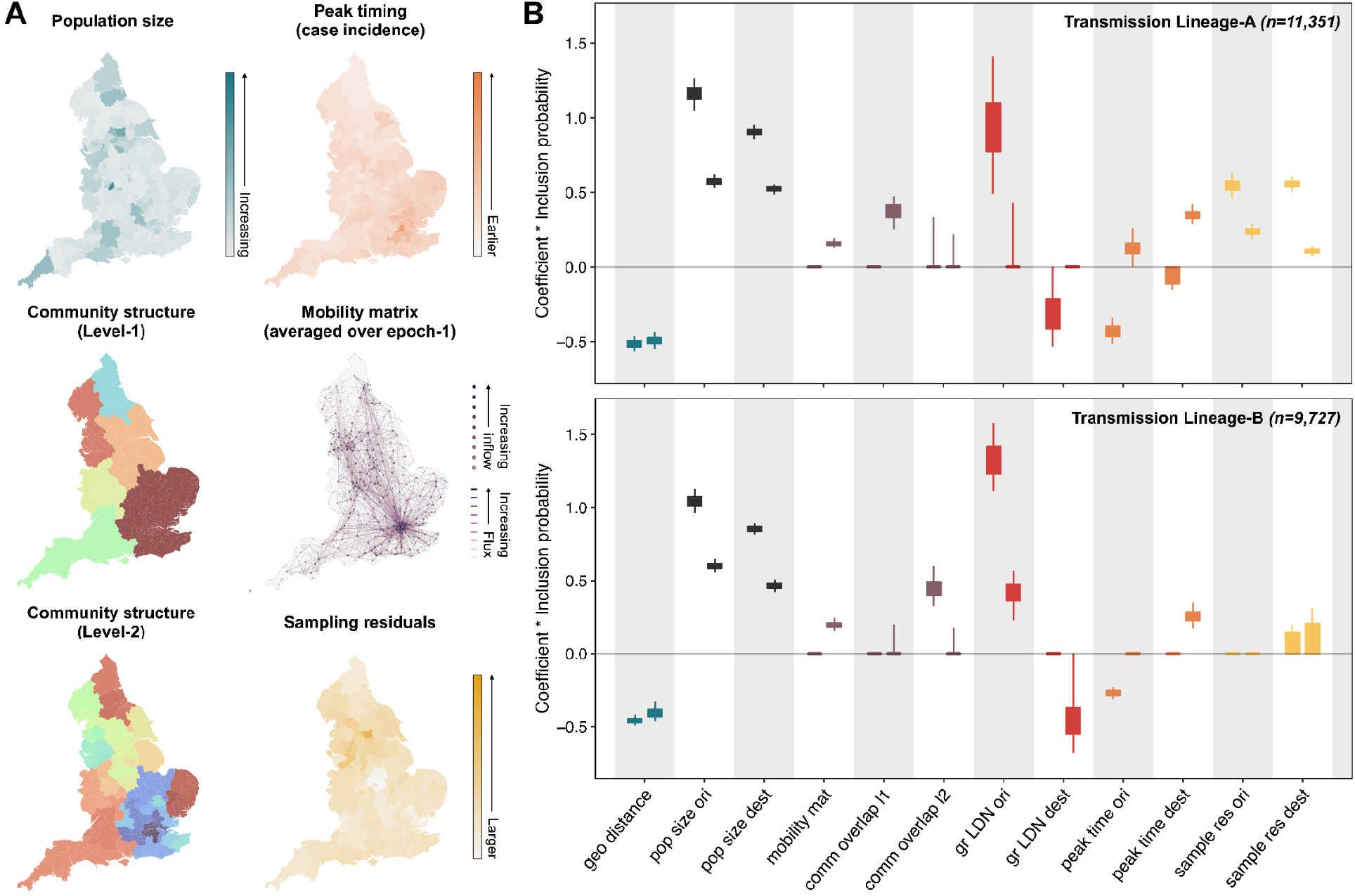
Predictors of Omicron BA.1 viral lineage movements in England. (A) Map at LTLA level of predictors considered in the discrete phylogeographic GLM analysis, for the largest detected BA.1 transmission lineages in England (Transmission Lineage-A). (B) Posterior distributions of the product of the log predictor coefficient and the predictor inclusion probability for each predictor. Estimates are plotted for the two largest transmission lineages: Transmission Lineage-A (top) and Transmission Lineage-B (bottom). For each predictor, the left hand value represents the expansion phase, the right hand value the post-expansion phase. Posterior distributions are coloured according to predictor type. Predictors include geographic distances (geo distance, red), population sizes at the origin and destination (pop size ori & pop size dest, yellow), aggregated mobility (mobility mat, dark blue), mobility-based community membership level 1 and level 2 (comm overlap l1 & l2, light blue), Greater London origin and destination (gr LDN ori & gr LDN dest, green), peak time at the origin and destination (peak time ori & peak time dest, yellow) and the residual of a regression of sample size against case count regression at either the origin and destination (sample res ori & sample res dest, grey).

**Figure 5:**
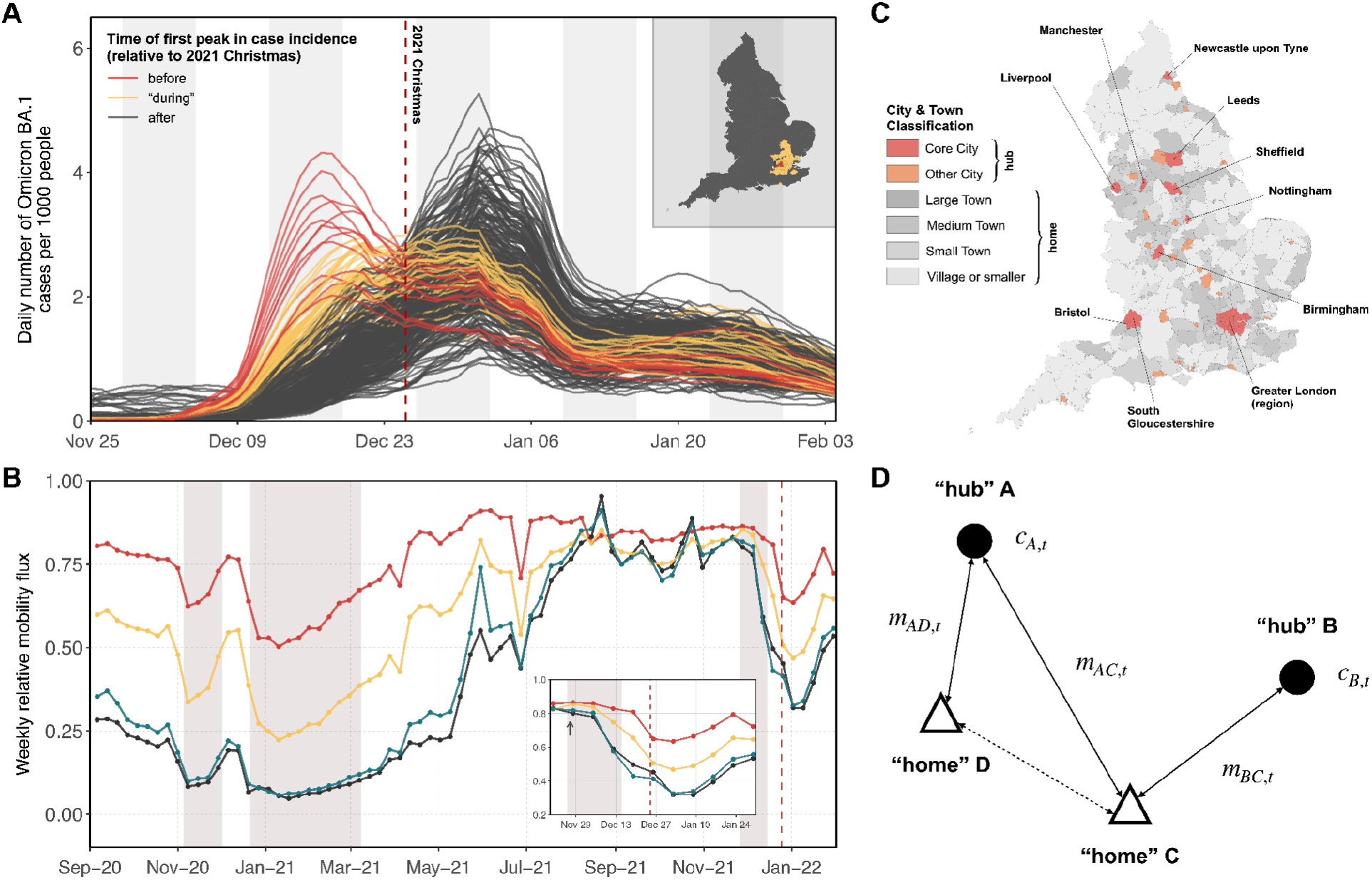
Spatiotemporal distribution of Omicron BA.1 infections and potential links with human mobility. (A) Estimated daily number of Omicron BA.1 cases per 1000 people at the LTLA level, coloured according to their timing of peak in case incidence (specifically, whether the interval during which the daily number of Omicron BA.1 cases exceeded 85% of the peak incidence lies entirely before (red), after (dark-grey), or includes (yellow) 2021 Christmas (25 December 2021). Inset shows the spatial distribution of LTLAs with different timing of peak in Omicron BA.1 case incidence. (B) Trend in human mobility in the UK across multiple spatial scales (red: <30 km, yellow: 30 - 60 km, green: 60 - 100 km, black: >100 km); arrow in inset indicates the time of first reported Omicron BA.1 infection in England on 27 November 2021. (C) Map showing the spatial distribution of LTLAs that are classified as one of 6 types of population settlement according to the City & Town Classification of Constituencies & Local Authorities, developed by the House of Common Library in the United Kingdom. (D) Schematic showing a mobility network with two “hub” units (A, B) and two “home” units (D, C). m_XY,t_ denotes the mobility flux between a “hub” unit X and “home” unit Y during a given week t, and c_X,t_ denotes the number of reported cases at “hub” X in a given week t. Only “home” unit C would be considered in the model in this example as unit D is only connected to a single “hub”.

Because the expansion dynamics are highly anisotropic, they offer little opportunity to find consistent support for the aggregated mobility predictor, which is symmetric. In contrast, the gravity model dynamics during the post-expansion phase(Figure 4B; Figure S7) are somewhat less pronounced, as suggested by consistently lower contribution from the population size predictor. Consequently, the aggregated mobility matrix is now consistently supported. For some transmission lineages, the model still suggests substantial viral lineage outflow from Greater London (i.e., support for Greater London predictors). In contrast to the initial expansion phase, the post-expansion phase exhibits consistently positive coefficients for the destination peak time predictor, reflecting higher dissemination to LTLAs with later peak times. Overall, these results are consistent with observations from continuous phylogeography (Figure 3) and epidemiological data from previous studies (*14, 15*). We further verify these results by performing a similar analysis using a time-homogeneous model, from which we find broadly consistent coefficient estimates for all predictors as those from the time-inhomogeneous model (Figure S8).

### Impact of prevalence on human behaviour

The apparent spatial heterogeneity in seeding and local establishment of BA.1 revealed in the phylogeographic analysis is also reflected in the case incidence data. Pre-holiday peaks were observed in Greater London and surrounding LTLAs (n=14) whereas most other LTLAs had BA.1 epidemic peaks in early January (n=255) (Figure 5A and B). Substantial reduction (∼20-50%) in human mobility across all spatial scales from mid-December 2021 until the first week of January 2022 might have partly contributed to the observed twin-peak distribution in epidemic curves (Figure 5A).

Changes in human mobility during this time can be attributed to three main factors: i) implementation of “Plan B” restrictions which included a work-from-home recommendation leading to reductions in commuting (*18*), ii) reduced mixing during the Christmas and New Year holidays, and iii) behavioural changes in response to increases in reported case numbers. Since i) and ii) were universally applied across all LTLAs in England, our subsequent analysis focuses on estimating the impact of heterogeneity in case incidence in major urban areas on the population’s behaviour.

Using a statistical model we estimate the impact of changes in the number of non-variant-specific COVID-19 cases per capita in a “hub” LTLA on the level of human mobility to/from a “home” LTLA (measured as the total amount of mobility flux regardless of direction). An LTLA is considered to be a “hub” unit if it is classified as a “Core City” or “Other City” by the City & Town Classification of Constituencies & Local Authorities (Figure 5C; Materials and Methods), or a “home” unit otherwise. By comparing only “home-hub” pairs involving the same “home” unit and assuming that these connections shared the same seasonal time trend (Figure 5D; Figure S9) (see Materials and Methods for further details), we find that the number of trips taken between a “home” and “hub” unit during the initial growth phase of the epidemic decreased by 11.97% [95% CI 2.84 to 20.81] for every unit increase in the weekly number of COVID-19 cases per 100 people at the “hub” in the same week (with a 4-day lag). We also consider geographical distance as an effect modifier and observe that longer distance movements were less affected compared to shorter distance movements; the percent reduction in connectivity for every unit increase in weekly cases per 100 people at the “hub” is estimated to be 20.03% [95% CI 14.50 to 25.18] and 15.79 [95% CI 10.29 to 20.94] when the “home” and “hub” units are separated by a distance of 50 km and 100 km respectively (Table S5). Whilst our results indicate that behavioural response to the growing epidemic was likely to have played a part in determining the velocity of spatial expansion of the Omicron wave in England, future studies involving detailed modelling and comparisons among different VOC waves in England are needed to fully disentangle the relative contribution from the three aforementioned factors.

## Discussion

Our results show that a substantial proportion of local SARS-CoV-2 infections during the Omicron BA.1 epidemic wave in England can be traced back to a small number of introductions inferred to have occurred before or during the early travel restrictions (26 November to 15 December 2021 for incoming passengers from southern Africa). Although the rate of international imports continued to increase after mid-December, local onward transmission was observed only for a small proportion (∼25%) of imported cases that arrived in England after Christmas 2021. These results are consistent with previous investigations of VOCs in England and other countries (*27, 35*), highlighting the ineffectiveness of international travel restrictions when applied after the detection of local transmission, when they are applied to too few countries, and not in combination with local control measures. This is especially true when countries outside the presumed origin location have rapidly growing epidemics and become major exporters of the VOC, as highlighted by the epidemic of BA.1 in NYC and continued exponential growth of importations into England in early December 2021 (*36*). More generally, the practical value of targeted travel restrictions is further limited by the fact that there are often multiple pathways connecting any two countries in the global transportation network (*37*).

Omicron BA.1 spread rapidly across England, with Greater London playing a central role in the initial spatial expansion due to its unique position as a central travel hub in England’s mobility network. In contrast, Greater London played a smaller role during the spread of Delta in Spring 2021 when most early transmission clusters were in the North West region, likely as a result of specific mobility patterns associated with incoming travellers (*17*). Similar spatial expansion patterns centred on Greater London were also observed in invasion of the Alpha variant in winter 2020 (*16*); future comparative studies of the dispersal patterns of local transmission lineages across these previous waves could potentially yield insights into the impact of NPIs and the effects of resulting changes of local mobility patterns on the spatial heterogeneity in the spread of VOCs and future infectious diseases.

Findings from our phylodynamic analysis should be interpreted in the context of several limitations. The number of seeding events identified is likely to be an underestimate of the true number of independent introductions due to unsampled genetic diversity outside of England as well as uneven sequencing coverage among countries (*38*). Some transmission lineages may have been aggregated, as indicated by extended periods of unobserved local transmission after the inferred importation. Nevertheless, we were able to cross-validate our phylogenetic results using independent epidemiological data from the VAM line list (Figure S6).

To remove potential sampling bias in the phylogeographic reconstruction of local transmission lineages, we downsampled the available English SARS-CoV-2 genomes from COG-UK whilst accounting for geographical variations in sequencing coverage and COVID-19 prevalence. Despite this subsampling procedure, we note that some sampling biases still remained both spatially (Figure 4A) and temporally (Figure S4). This could be due to geographical variations in case reporting rate or because the maximum sequencing capacity was exceeded in regions with exceptionally high case incidence. Our results indicate evidence of voluntary adjustments in human mobility patterns in response to changes in the number of Omicron BA.1 cases reported locally and at major local travel hubs.

Future work should focus on evaluating how the level of coupling between intensity of transmission and human behaviours varies depending on socio-economic factors and on levels of NPIs or risk communication. For example, regions or countries where most of the workforce is employed in manual labour are unlikely to exhibit the same voluntary behavioural response as those where more jobs can be performed from home (*39, 40*), thus allowing employees more freedom to respond to local changes in prevalence. Similarly, individuals with different socio-economic status are likely to display varying degrees of behaviours due to differences in exposure to health information (*41, 42*).

Understanding the bi-directional relationships between human behaviours and perceived level of transmission risk is crucial for the design of future intervention strategies and modelling for COVID-19 and other respiratory infections. Additionally, they should also explore how spatial heterogeneities in population immunity may impact SARS-CoV-2 spread.

Our findings highlight the need to better characterise the interplay between perceived risk and intensity of transmission and changes in human behaviour. Their interaction dynamics and the resulting impact on transmission could be relevant to the way in which many countries, including the UK, move toward managing endemic COVID-19. Omicron BA.1 was replaced by the BA.2 sub-variant in February 2022 and later by the BA.5 sub-variant in June 2022 (*20, 21*). Whilst the public health burden of COVID-19 has remained lower due to reduced average disease severity and increased population immunity, the continued antigenic evolution of SARS-CoV-2 means that the possibility of future variants with increased virulence cannot be dismissed. One priority in preparing for the next SARS-CoV-2 variant or pathogen emergence is developing robust and scalable pipelines for large-scale genomic and epidemiological analyses supported by globally coordinated and unified data infrastructures (*43, 44*). The implementation of analytical systems which draw on multiple data streams comes with myriad technical and logistical challenges and can be realised only through the collective efforts of public health stakeholders worldwide.

## Data Availability

UK genome sequences used were generated by the COVID-19 Genomics UK consortium (COG-UK, https://www.cogconsortium.uk/). Data linking COG-IDs to location have been removed to protect privacy, however if you require this data please visit https://www.cogconsortium.uk/contact/ for information on accessing consortium-only data. The Google COVID-19 Aggregated Mobility Research Dataset used for this study is available with permission from Google LLC. Code to reproduce the analyses will be made available on our GitHub repository before publication.

## Acknowledgements

COG-UK is supported by funding from the Medical Research Council (MRC) part of UK Research & Innovation (UKRI), the National Institute of Health Research (NIHR) [grant code: MC_PC_19027], and Genome Research Limited, operating as the Wellcome Sanger Institute. The authors acknowledge use of data generated through the COVID-19 Genomics Programme funded by the Department of Health and Social Care. MUGK acknowledges funding from The Rockefeller Foundation, Google.org, the Oxford Martin School Pandemic Genomics programme (also O.G.P. and A.E.Z.), European Union Horizon 2020 project MOOD (#874850) (also supports R.I., S.D. and P.L.), The John Fell Fund, and a Branco Weiss Fellowship. This work was also supported by the Foreign, Commonwealth & Development Office and Wellcome [225288/Z/22/Z] and [226052/Z/22/Z] (to M.U.G.K.). V.H. was supported by the Biotechnology and Biological Sciences Research Council (BBSRC) [grant number BB/M010996/1]. J.T.M, R.C. and A.R. acknowledge support from the Wellcome Trust [Collaborators Award 206298/Z/17/Z - ARTIC network]. SD also acknowledges support from the *Fonds National de la Recherche Scientifique* (F.R.S.-FNRS, Belgium; grant n°F.4515.22) and from the Research Foundation – Flanders (*Fonds voor Wetenschappelijk Onderzoek* – *Vlaanderen*, FWO, Belgium; grant n°G098321N). A.R. is also supported by the European Research Council [grant agreement number 725422 - ReservoirDOCS] and Bill & Melinda Gates Foundation [OPP1175094 – HIV-PANGEA II]. SB is supported by the Clarendon Scholarship, University of Oxford and NERC DTP [grant number NE/S007474/1].]. M.A.S. acknowledges support from US National Institutes of Health grants R01 AI153044 and R01 AI162611. M.A.S. and X.J. gratefully acknowledge support from NVIDIA Corporation and Advanced Micro Devices, Inc. with the donation of parallel computing resources used for this research. SC acknowledges Labex IBEID (grant ANR-10-LABX-62-IBEID), European Union Horizon 2020 projects VEO (874735) and RECOVER (101003589), AXARF, Groupama, EMERGEN (ANRS0151) and INCEPTION (PIA/ANR-16-CONV-0005). E.W., H.T. and T.dO are supported in part by grants from the Rockefeller Foundation (HTH 017), the Abbott Pandemic Defense Coalition (APDC), the African Society for Laboratory Medicine, the National Institute of Health USA (U01 AI151698) for the United World Antivirus Research Network (UWARN) and the INFORM Africa project through IHVN (U54 TW012041)The views expressed are those of the author and not necessarily those of the Department of Health and Social Care, UKHSA, or European commission or any of the other funders.

## Data Acknowledgements

We acknowledge the UK Health Security Agency (UKHSA), members of the COVID-19 Genomics UK (COG-UK) consortium, NHS labs, GISAID contributors (acknowledgment table of genomes used is provided on our GitHub repository) for sharing of genomic data of Omicron BA.1.

## Author contributions

JT, OGP, MUGK conceived and planned the research. JT, SB, BL,VH, JTM, PB, REP, JR, PL, SD analysed the data. AEZ, TPP, BJ, RC, AO’T, AD, JTM, BL, SC, SD, JR, XJ, MAS, MUGK, OGP, AR advised on methodologies. IIB, KK, MD contributed international flight passenger data. JT, OGP, MUGK wrote the initial manuscript draft. All authors edited, read and approved the manuscript.

## Materials and Methods

### Genomic data

All SARS-CoV-2 sequences used in this study were downloaded on 12 April 2022. All available international (non-England, i.e. including Wales, Scotland and Northern Ireland) sequences were downloaded from GISAID while English samples marked as community surveillance (pillar 2) were acquired from COG-UK. Historically, pillar 2 testing sites were instructed to select a number of 96 well plates for sequencing proportional to the fraction of total tests that week. Pillar 2 surveillance is intended to represent a random sample of community cases in the UK; however, given the changes in testing behaviour and regulations that occurred during the study period we can not rule out the possibility that there are some biases in the data set. These were partially addressed in the subsampling mentioned below.

Sequences were aligned and filtered as part of the COG-UK datapipe analysis hosted by CLIMB. This analysis removed duplicate and environmental sequences, and flagged samples with improbable collection dates (see https://github.com/COG-UK/datapipe for details). All sequences with impossible or improbable collection dates were removed. To further minimise dating errors caused by retrospective sequencing, only samples published to COG-UK or GISAID within four weeks of sample collection were included. Sequences were aligned to the reference Wuhan-Hu-1 (genbank accession MN908947.3) with minimap2 and samples with less than 93% coverage were discarded.

Sequence coverage weights were calculated for English sequences (https://github.com/robj411/sequencing_coverage) to ensure they could be subsampled proportional to the number of reported cases in each Upper Tier Local Authority using a two week sliding window.

Scorpio was run as part of Pangolin and sequences identified as BA.1 or BA.2 were selected for further analysis.

### Variant and Mutations (VAM) line list (from UK Health Security Agency)

The variant and mutations (VAM) line list compiled and provided by the UK Health Security Agency (UKHSA) contains epidemiological metadata of specimens sequenced from the pillar 2 mass testing programme by the COVID-19 Genomics UK Consortium (COG-UK). From the variant line list we extracted the traveller status (Traveller, Contact of Traveller, Not Travel-associated, Refused or Uncontactable, Awaiting Information) and specimen date of all sampled individuals who were tested positive for Omicron B.1.1.529 (BA.1) between 1 November 2021 and 31 January 2022.

### Estimated Omicron BA.1 case incidence (from COVID-19 case count and S-gene target failure data)

Daily number of new COVID-19 cases by specimen date in each LTLA were downloaded from https://coronavirus.data.gov.uk/details/download (last assessed on 26 June 2022). S-gene target failure data were provided by UKHSA via a data sharing agreement. The presence of a genetic deletion on the spike protein of the Omicron BA.1 sub-variant produces SGTG in most PCR tests which can be used as a proxy for BA.1 infections. We used daily SGTF PCR-positive tests as a proxy (because these were time-and cost-effective as a test compared to genetic sequencing to ascertain variants) for Omicron BA.1 infection in conjunction with reported case data to estimate daily number of new BA.1 cases. However, small sample sizes in the SGTF dataset could lead to extreme scaling, i.e. zero or 100% of cases could be attributed to BA.1 infections if for example none or all samples were SGTF positive. Hence we calculated BA.1 cases in a Bayesian framework using uninformative Beta(1, 1) priors and the observed proportion of BA.1 infections (from the SGTF dataset) to estimate the posterior proportion of BA.1 cases which was then scaled up by the number of reported cases from the coronavirus data download. We can also use the estimated uncertainty from the posterior distribution to get lower and upper bounds in the scaled up BA.1 case numbers.

### International passenger flight data for South Africa and the United States

We evaluated travel data generated from the International Air Transport Association (IATA) to quantify passenger volumes originating from international airports in South Africa and the United States. IATA data accounts for approximately 90% of passenger travel itineraries on commercial flights, excluding transportation via unscheduled charter flights (the remainder is modelled using market intelligence).

### Estimated importation intensity of Omicron BA.1 from South Africa and the United States

We estimated and compared the weekly importation intensity of SARS-CoV-2 Omicron BA.1 from South Africa and the United States between 7 Nov 2021 and 26 March 2022. The weekly importation intensity is an estimate of the number of Omicron BA.1 cases imported into the UK during a given week from a specified source location, calculated by multiplying together the estimated weekly prevalence of the Omicron BA.1 variant at the source location and the number of air passengers arriving in the UK from the source location.

To obtain an estimate of the weekly prevalence of Omicron BA.1 at the source locations, we multiplied the weekly number of reported COVID-19 cases per capita with the proportion of sequenced SARS-CoV-2 genomes that were of the Omicron BA.1 variant in the same week (as available from GISAID, https://gisaid.org/; last accessed on 21 April 2022). Data on the number of reported COVID-19 cases per week were downloaded from OWID (https://ourworldindata.org/; last accessed on 9 Nov 2022) and The New York Times (https://github.com/nytimes/covid-19-data; last accessed on 13 Dec 2022) for South Africa and the US respectively. For the US, weekly case counts per capita are available only at the state-level, which we assumed to be homogeneous and therefore representative of the weekly case counts per capita in individual cities in the same states.

To obtain an estimate of the weekly number of air passengers arriving from the US in the UK, we considered only the top 20 cities with the most intense incoming traffic (between October 2021 to March 2022; collectively accounting for ∼87% of air traffic from the US to the UK). For South Africa, air passenger data is only available at the country-level and at monthly intervals. To estimate the weekly numbers of air passengers arriving from South Africa, we assumed an even daily distribution of passengers throughout the month which we then aggregated at the week-level.

### UK population data

Mid-year population estimates for England in 2020 at the LTLA level were downloaded from https://www.ons.gov.uk/peoplepopulationandcommunity/populationandmigration/populationestimates/datasets/populationestimatesforukenglandandwalesscotlandandnorthernireland. Population sizes were used as the denominator in calculating numbers of COVID-19 cases per capita and normalised local mobility in each LTLA.

### Aggregated and anonymised human mobility data

We used the Google COVID-19 Aggregated Mobility Research Dataset described in detail in (*45, 46*), which contains anonymized relative mobility flows aggregated over users who have turned on the *Location History* setting, which is turned off by default. This is similar to the data used to show how busy certain types of places are in Google Maps — helping identify when a local business tends to be the most crowded. The mobility flux is aggregated per week, between pairs of approximately 5km^2^ cells worldwide, and for the purpose of this study further aggregated for LTLAs in the United Kingdom (https://geoportal.statistics.gov.uk/datasets/lower-tier-local-authority-to-upper-tier-local-authority-december-2016-lookup-in-england-and-wales/explore) for the time period of November 2019 to January 31st, 2022.

To produce this dataset, machine learning is applied to log data to automatically segment it into semantic trips. To provide strong privacy guarantees (*47*), all trips were anonymized and aggregated using a differentially private mechanism to aggregate flows over time (see https://policies.google.com/technologies/anonymization). This research is done on the resulting heavily aggregated and differentially private data. No individual user data was ever manually inspected, only heavily aggregated flows of large populations were handled. All anonymized trips are processed in aggregate to extract their origin and destination location and time. For example, if n users travelled from location a to location b within time interval t, the corresponding cell (a,b,t) in the tensor would be n∓err, where err is Laplacian noise. The automated Laplace mechanism adds random noise drawn from a zero mean Laplacian distribution and yields (∈, δ)-differential privacy guarantee of ε = 0.66 and δ = 2.1 × 10−29 per metric. Specifically, for each week W and each location pair (A,B), we compute the number of unique users who took a trip from location A to location B during week W. To each of these metrics, we add Laplace noise from a zero-mean distribution of scale 1/0.66. We then remove all metrics for which the noisy number of users is lower than 100, following the process described in (*47*), and publish the rest. This yields that each metric we publish satisfies (ε,δ)-differential privacy with values defined above. The parameter ε controls the noise intensity in terms of its variance, while δ represents the deviation from pure ε-privacy. The closer they are to zero, the stronger the privacy guarantees.

These results should be interpreted in light of several important limitations. First, the Google mobility data is limited to smartphone users who have opted in to Google’s *Location History* feature, which is off by default. These data may not be representative of the population as whole, and furthermore their representativeness may vary by location. Importantly, these limited data are only viewed through the lens of differential privacy algorithms, specifically designed to protect user anonymity and obscure fine detail. Moreover, comparisons across rather than within locations are only descriptive since these regions can differ in substantial ways.

### Phylogenetic analyses

#### Importation analysis

We developed a large-scale phylogenetic analysis pipeline following a similar approach as in du Plessis et al. (2021) (*48*) with additional extensions and modifications to ensure the computational tractability of analyses of up to hundreds of thousands of SARS-CoV-2 sequences (*49*).

First, the study period was divided into two phases: (1) from 21 November 2021 (sample date of the earliest known genome of the Omicron variant in England, sequenced retrospectively) to 28 November 2021, and (2) from 29 November 2021 to 31 January 2022. The time of division between the two phases was chosen on the basis of an expected change in importation intensity as a result of the implementation of travel restrictions targeted at multiple southern African countries starting on 28 November 2021. With the relatively few genomes available from the first phase and to account for an increased risk of importations prior to the travel restrictions, all 874 available sequences (from both England and non-England locations) were included. Owing to the large number of genome samples collected during the second phase, a downsampling strategy was applied to ensure that the analysis was computationally tractable. To generate a manageable dataset of global sequences, first we computed a crude estimate of the number of new Omicron cases in each country in each epi-week by multiplying the number of reported COVID-19 cases (downloaded from https://github.com/owid; last accessed on 4 May 2022) by the proportion of sampled genomes that were of the Omicron variant PANGO lineages BA.1 and BA.2, using metadata available from GISAID (https://gisaid.org/; last accessed on 12 April 2022). The number of global sequences to be sampled in each epi-week was then allocated in proportion to the estimated total number of Omicron lineages BA.1 and BA.2 cases in the week whilst maintaining a dataset size of ∼50,000. In a given epi-week, countries with an estimated number of Omicron cases that accounted for at least 0.5% of the estimated global total were considered as potential exporters. Genome samples were then allocated in proportion to the estimated number of cases among these potential exporters, with the remaining allocation randomly distributed among the non-exporter countries. There was a slight enrichment for samples collected in the early phase of the first Omicron wave (early December 2021), where we ensured that a minimum of 4,000 genomes were sampled for each epi-week where available. A similar approach was used to curate a dataset of 21,039 Omicron genomes sampled from Wales, Scotland and Northern Ireland, again using relevant metadata from GISAID and epidemiological data available on (https://api.coronavirus.data.gov.uk/v1/data; last accessed on last accessed on 4 May 2022). This downsampling procedure resulted in a dataset of 59,647 global (non-English) sequences. To generate a dataset of English genomes of roughly the same size, 60,000 sequences were randomly sampled from the COG-UK master alignment whilst accounting for variations in sequencing coverage and prevalence amongst UTLAs over time, using the same method as in Volz et al. (*50*). This resulted in a combined dataset of 140,686 genomes of which 42.6% were sampled in England with the remaining from non-England locations.

Despite substantial downsampling, estimating a phylogenetic tree for hundreds of thousands of SARS-CoV-2 sequences remains a challenge, with most standard programs only able to handle up to thousands of sequences. To tackle this, we first estimated a maximum likelihood (ML) tree for the 874 sequences collected during the first phase of the study period using IQTREE (*51*) with the GTR+G substitution model, rooted with reference genome Wuhan-Hu-1 (GenBank accession MN908947.3) as an outgroup. Five molecular clock outliers were identified and subsequently removed, after examining the root-to-tip regression plot from TreeTime (*52*). The resulting tree was then used as a starting tree from which a parsimony tree was estimated by inserting individual sequences sequentially and in chronological order according to sample dates, using the recently developed UShER placement tool (*53*). During each step in the iterative process, all sequences sampled on a given date were considered for placement whilst excluding sequences with 5 or more equally parsimonious placements.

Sequences excluded in a previous step were appended to the next batch for reconsideration. The resulting tree was then optimised through 6 iterations of matOptimize (*54*) with SPR radius of 40 and 100 for the first 5 and final iteration respectively. This iterative tree building process resulted in a phylogeny of 115,634 sequences (with 25,921 (18.3%) sequences excluded due to uncertainty in sample placement). Next we used Chronumental (*55*) (a recently developed time-tree estimation tool for handling large phylogenies) to estimate a randomly resolved time-calibrated tree, with inferred tip dates that maximise the evidence lower bound under a probabilistic model. By comparing the inferred tip dates with sample dates and examining a root-to-tip plot, 12 molecular clock outliers were further removed, resulting in a final phylogeny of 115,622 sequences.

To further reduce the computational resources and time required, we divided the phylogeny estimated above into smaller tree partitions according to sub-lineage (of Omicron) assignment as defined by the PANGO nomenclature (*56*). Using a custom Python script, subtrees with a high degree of clustering of sequences of the same descendant lineage of Omicron were identified, whilst accounting for some level of ambiguity in lineage assignment (e.g. a tree partition may contain up to 25% of sequences that are of a minority sub-lineage before it is subdivided into multiple partitions), as would be expected given the high sampling density and variations in sequencing quality. Further merging of these identified subtrees resulted in five final tree partitions, labelled BA.1 (n=38,522), BA.1.1 (n=37,028), BA.1.15 (n=12,229), BA.1.17 (n=21,549), and BA.2 (6,294) according to the sub-lineage represented by the majority of sequences in each partition. Given that the primary focus of this study is the invasion dynamics of Omicron BA.1 in England, the BA.2 partition was omitted in all further downstream analyses.

Having divided the phylogeny into smaller tree partitions of computationally manageable size, we then performed time-calibration of the subtrees using a recently implemented model in BEAST v1.10 (*57*) which replaces the traditional tree-likelihood with a more efficient likelihood based on a simple Poisson model, thus allowing Bayesian phylogenetic analyses of up to tens of thousands of sequences. In this approach, the operators are constrained such that only node heights and polytomy resolutions are sampled, whilst the tree topology is fixed to that of a data tree which we generated using Treetime (*52*) with a fixed clock rate of 7.5×10E-4 substitutions/site/yr. Using a Skygrid coalescent tree prior (*58*) with grid points every two weeks, we ran between 2 and 6 MCMC chains of 3×10^8^ to 2.4×10^9^ iterations for each tree partition independently. The first 33% to 40% of each chain was discarded as burn-in and resampled every 1×10^6^ to 2.4×10^8^ states before merging using LogCombiner, resulting in 1,200 posterior tree samples for each tree partition. Model convergence and mixing was assessed using Tracer (*59*).

To reconstruct the importation dynamics of Omicron BA.1, we then used a two-state asymmetric discrete trait analysis (DTA) model implemented in BEAST v1.10 (*57*), using the posterior tree samples estimated above as the empirical tree distributions. For each tree partition, we ran two MCMC chains of 5 million iterations each, resampled every 9,000 states and with the first 10% discarded as burn-in. TreeAnnotator 1.10 (*57*) was used to generate a maximum clade credibility (MCC) tree for each subtree, in which each internal node is assigned a posterior probability of representing a transmission event in England. Nodes with a posterior probability of >0.5 were identified as introductions; a small number of nodes with ambiguous location assignment (posterior probability = 0.5) were ignored in downstream analyses. To identify the local transmission lineage resulting from each of the introductions, a depth-first search was performed following the same procedure as in du Plessis et al. (2021) (*48*), where a path starting from each internal node that corresponds to an introduction is traversed forwards in time until a non-England node is encountered or there are no more nodes to be explored. By convention, introductions that led to only a single sampled English sequence were labelled as singletons; only introductions that led to more than one observed local transmission event were labelled as transmission lineages. The time of importation of each transmission lineage was estimated by taking the mid-point between the internal node corresponding to the introduction and its parent.

Our methodology estimating the time of importation of transmission lineages is likely to result in an apparent “expansion” of the temporal profile of inferred importation intensity (daily number of infected travellers arriving in England) relative to its true underlying distribution. This could be explained by an increase in importation lag (time elapsed between when a lineage is inferred to have been imported and the first detected local transmission event) over time as shown previously by du Plessis et. al. (*27*), due to transmission lineages from later importations having fewer genomes as they have less time to grow, and are therefore less likely to be captured by genomic surveillance assuming a constant sampling intensity. To verify this effect, we compared the inferred importation intensity from the above phylodynamic analysis with empirical observations from testing data recording international travellers, extracted from the Variant Mutation line list provided by UKHSA (Figure S1). However, we note that the robustness of this comparison is potentially limited by variations in sampling intensity in the epidemiological data as a result of rapidly-changing testing policies for arriving travellers in the United Kingdom during the latter part of January 2022 (*60*).

#### Exponential growth of daily frequency of importations

In the absence of any travel restrictions and changes in human mobility as a result of the emergence of a new VOC, the importation intensity during the initial phase of the invasion would be expected to follow a pattern of exponential growth that mirrors the increase in number of infections in the exporting countries. To verify and examine any potential deviation from this pattern, we fitted a simple exponential model to the 7-day rolling average daily number of importations inferred from the phylodynamic analysis. Specifically, we fitted the model using least-squares regression to the inferred daily numbers of importations during the period between the beginning of November 2021 and a range of cut-off dates. The cut-off date that resulted in the highest adjusted R^2^ value can be interpreted as an estimate of the time when the growth of importation intensity began to deviate from an exponential trajectory.

#### Continuous phylogeography

To reconstruct the spatiotemporal patterns of the Omicron BA.1 wave in England, all local transmission lineages (as identified from the MCC trees generated from the 2-state discrete trait analysis above) with five or more sequences were extracted for continuous phylogeographic analyses. Each sequence was assigned a latitude and a longitude randomly from within the postal district (metadata provided by COG-UK) where the sample was collected. For each transmission lineage, we performed the continuous phylogeographic reconstruction on a fixed (pruned from the MCC tree) using a relaxed random walk model (*61*) implemented in BEAST 1.10.4 (*57*), with a Cauchy distribution to model the among-branch heterogeneity in dispersal velocity. Following a similar approach as in McCrone et al. (*17*), the 8 largest transmission lineages (labelled A to H, from largest to smallest) containing >700 sequences were inferred independently, with the remaining smaller transmission lineages (n=524) inferred in a single joint analysis with a shared diffusion model (i.e. same parameter estimates for likelihood, precision matrix, correlation, etc, but independent estimates for diffusion rate and trait likelihood). Owing to variations in the extent of spatial dispersal among these smaller transmission lineages (with larger lineages being more spatially dispersed in general), 30 were subsequently removed from the joint analysis and inferred independently. Model convergence and mixing was assessed using Tracer v1.7 (*59*). For the independent analyses of the 8 largest transmission lineages, we ran between 2 and 5 MCMC chains of 200 to 300 million iterations, sampling every 10,000 to 80,000 states and removing the first 10% to 33% of each chain as burn-in, resulting in 10,000 to 13,5000 trees sampled from the posterior distribution. For the independent analyses of the 30 smaller transmission lineages with fewer than 700 sequences, we ran 2 MCMC chains each of 200 million iterations which we then merged after resampling every 30,000 states and removing the first 10% as burn-in, giving 12,000 posterior trees per transmission lineage. Finally, in the joint analysis, 8 independent chains of 200 million were run with sampling every 120,000 states. They were combined after removing the first 10% as burn-in, again resulting in 12,000 posterior tree samples for each transmission lineage. These posterior tree samples were then used to generate an annotated MCC tree for each transmission lineage using TreeAnnotator (*57*).

To facilitate subsequent analyses of viral lineage movements at the LTLA level, we mapped the inferred location of each internal node in the transmission lineages to its corresponding LTLA by checking whether the inferred coordinates are contained within the associated polygon. In the case where an enclosing polygon could not be found (e.g. a small proportion of internal nodes were inferred to lie in the small spaces between neighbouring polygons), the polygon that is geographically closest to the inferred location was assigned.

#### Discrete phylogeography with generalised linear model (GLM) parameterisation

We used the approach of discrete phylogeography with generalised linear model (GLM) to parameterise transition rates between locations and test the association of viral lineage dispersal with a number of geographical, demographic, epidemiological and human mobility-related factors (see Table S2 for full list of predictors). Specifically, to test the gravity model as a predictor of viral lineage movements, we considered in the GLM analysis the population size at the origin and destination location of each movement and the geographical distance between them. To further capture any heterogeneities in aggregated human mobility at the city-level (which are unlikely to be adequately described by the gravity model), we also included the aggregated mobility matrix and community memberships as predictors. We allowed these mobility-related predictors to vary across different phases in the time-inhomogeneous model to test for temporal variations in aggregated human mobility patterns and also potentially time-varying effect of mobility on viral dispersal. We observed from both epidemiological data and continuous phylogeography that many LTLAs in Greater London experienced an earlier uptick in Omicron BA.1 cases compared to most LTLAs with other regions of England. To capture this asynchronicity in local epidemic dynamics and investigate its impact on viral dispersal, we considered in the GLM analysis whether each viral movement started or ended in the Greater London region and additionally the time of first peak in Omicron BA.1 case incidence at the origin and destination location. Furthermore, we also tested for the impact of sampling bias by including a predictor based on the residuals from a simple regression of sample size against Omicron BA.1 cases for both the origin and destination location. Due to the small number of sequences collected in some LTLAs especially during later phases of the epidemic, the regression residuals were computed using sample sizes and case counts aggregated over the whole study period in both the time-homogeneous and time-inhomogeneous models.

Unlike continuous phylogeography where each sequence is assigned a unique set of coordinates in continuous space, discrete phylogeography requires that sequences are grouped into discrete geographical units. The level of granularity of these geographical units depends on a number of factors including (i) the desired level of resolution at which the dispersal history is to be reconstructed, (ii) the amount of heterogeneities present within each geographical unit, and (iii) the maximum number of geographical units beyond which the analysis becomes computationally intractable. To capture heterogeneities in viral movements at the city-level and to allow comparisons with results from continuous phylogeography, we allocated sequences to their corresponding lower tier local authorities (LTLA) using a lookup table which provides unique mapping between postal districts and LTLAs.

The current computational architecture and implementation of the discrete phytogeographic GLM model limits the number of discrete units possible to 256, which is smaller than the number of LTLAs across which sequences were sampled for some of the larger transmission lineages. To tackle this, we aggregated LTLAs where appropriate to reduce the number of geographic units. In order to minimise the resulting information loss, we first considered LTLAs with the fewest sequences and performed a merging operation if an adjacent LTLA with at least one sampled genome could be found. In the case where multiple adjacent LTLAs were available, the LTLA with the most number of sampled genomes was chosen for the merger. After each merging operation, the list of LTLAs (or geographical units after merging) ranked by the number of sampled genomes was recalculated for the next iterative step (it is therefore possible for an LTLA to be involved with multiple merging operations). This process continued until there were only 253 geographic units in each transmission lineage. For the geographical units consisting of multiple LTLAs, each statistic of interest was averaged over the relevant LTLAs, weighted by population size where appropriate. For transmission lineages with sequences sampled in fewer than 256 LTLAs, no merging was performed.

The discrete phylogeographic GLM model parameterizes the log of between-location transition rates as a log linear function of the predictors. Continuous predictors (geographical distances, population sizes, aggregated mobility matrices, peak timing in case incidence, sampling residuals) were therefore log-transformed and standardised after adding a pseudo-count to each entry where appropriate. Binary variables (community memberships, Greater London/non-Greater London) were encoded as 0 and 1. In the mobility-related predictors, there was missing data for one or two geographical units in some transmission lineages (due to mobility data being unavailable for South Tyneside and City of London), which we labelled as NA and later integrated out in our Bayesian inference. For the aggregated mobility matrix predictor with continuous values in the large-scale transmission analyses, we confronted this using a new Hamiltonian Monte Carlo (HMC) kernel to jointly sample all missing covariates from their posterior distributions building on similar efforts in the BEAST framework (*62, 63*). The HMC kernel produces distant proposals with relatively high acceptance rate for the Metropolis algorithm by exploiting numerical solutions to the Hamiltonian dynamics. We performed the analyses using the code available in the hmc-clock branch of the BEAST codebase (available at https://github.com/beast-dev/beast-mcmc/tree/hmc-clock) in conjunction with the BEAGLE code available in the hmc-clock branch of the codebase (available at https://github.com/beagle-dev/beagle-lib/tree/hmc-clock). We ran the analyses on a set of 100 empirical trees for each transmission lineage extracted from the BEAST importation analysis and ran sufficiently long chains sampling every 500 generations, or combined multiple chains (excluding adequate burn-ins), to ensure effective sample sizes (ESSs) > 100 for the continuous parameters as diagnosed using Tracer (*59*). A custom R script was used to summarise and visualise the posterior coefficient estimates and inclusion probabilities of each predictor.

#### Branching process model and comparison of transmission lineage size distributions

To verify that the time of importation is the key determinant of transmission lineage size, we compared the size distribution of empirically observed transmission lineages with that from a model that simulates the branching process of transmission lineages following importation. Simulated importation dates are set to the dates estimated from the phylodynamic analysis and simulated transmissions occur at the spatially homogeneous growth rates estimated from the daily number of reported COVID-19 cases and SGTF data in England. Due to the low number of Omicron BA.1 cases at the beginning of the epidemic, which can lead to unreliable estimates of the initial growth rate, we performed a series of simulations with a range of different starting growth rates (taken from estimates during early parts of the invasion). We computed the Kullback-Leibler (KL) distance between the size distribution of simulated lineages and that of lineages inferred from phylodynamic analysis (Table S1). The growth rate that minimised the KL distance was then used to impute the initial growth rate in the best-fit model.

#### Statistical and human mobility analyses

We modelled the changes in connectivity between two locations over time, in response to changes in the number of new COVID-19 cases per capita. We denote the connectivity between two locations *i* and *j* during a given week *t* by 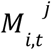, which we measure as the weekly total relative flow between *i* and *j* regardless of direction. Importantly, we make the assumption that the connectivity between two locations *i* and *j* only depended on the local characteristics at *i* and *j*, but not those at other connected locations in the mobility network. Whilst it is reasonable to expect that the number of trips taken between two locations during a given week should be most affected by changes in characteristics at the origin and destination, the possibility of transit journeys means that some of the changes in the mobility flow between the two locations in question may be attributable to changes in characteristics in a third connected location. We minimise this spillover effect in our following analyses by considering only connectivity between geographical units labelled as “home” and “hub”, such that the observed mobility flow between any pairs of “home” and “hub” units can be reasonably assumed to be mostly attributable to journeys taken between the two locations rather than transit journeys.

Given that the focus of this analysis is to assess the impact of changes in case incidence on human mobility patterns during the early growth of the epidemic, we defined the study period to be the five weeks following the first detection of Omcron in England (i.e. from the week commencing on 28 November 2021 to the week ending on 2 January 2022). Each statistic of interest was aggregated at the weekly level, with Sunday defined as the start of each week. To allow a lagged effect of COVID-19 cases on mobility, we introduced a 4-day delay in the time-windows over which case numbers were aggregated (i.e. the numbers of new COVID-19 cases per week were calculated with Wednesday as the start of each week).

The degree to which COVID-19 case incidence impacts human mobility is likely to depend on the nature and purpose of the movements. For example, human movements associated with social activities are likely to suffer a greater impact as a result of a change in case incidence compared to work-related travels. To estimate this effect, we considered the geographical distance between a “home” and “hub” unit as a proxy for the different types of movements which we included as an effect modifier in a subsequent model. In the case where multiple LTLAs have been aggregated into a “hub” unit, the new centroid of the combined polygons was used in calculating the geographical distances.

#### Classification of “home” and “hub” units

The House of Commons Library under the UK Parliament (downloaded from https://researchbriefings.files.parliament.uk/documents/CBP-8322/lauth-classification-csv.csv) developed a classification of constituency and local authority areas (LTLA) according to the type of settlement in which the largest proportion of its population lives, with the following 6 categories: “Core City” (e.g. London, Glasgow, Birmingham), “Other City” (e.g. Leicester, Coventry, Leeds), “Large Town”, “Medium Town”, “Small Town”, and “Village and small community”. Using this classification, each of the 314 LTLAs in England was labelled as either a “hub” unit if it is in the “Core City” or “Other City” category, or a “home” unit otherwise. Due to changes to certain LTLA definitions since the publication of the City & Town Classification in 2018, some deprecated LTLAs (with potentially different City & Town assignments) have been aggregated into larger units according to the most recent definitions (see Table S4 for further details). In such cases, these newly defined composite LTLAs were labelled as “hub” units if they contain at least one deprecated LTLA that was in the “Core City” or “Other City” category. Finally, in the case where multiple adjacent LTLAs were labelled as “hub” units (e.g. in Greater London), they were aggregated and considered as a single “hub” unit in all subsequent analyses. This procedure resulted in a final selection of 65 LTLAs, of which 40 were further aggregated to give 30 “hub” units.

According to the City & Town Classification, 248 LTLAs were not in the “Core City” or “Other City” category. An important further criterion for one of these LTLAs to be further considered as a “home” unit is that it must be connected to at least two “hub” units throughout the study period, thus excluding any LTLAs that were in the peripheral parts of the mobility network and were only connected to the “hub” units through other “home” units. This selection criterion resulted in 105 LTLAs being selected as “home” units (see Table S3 for list of all “home” and “hub” units).

#### Main regression

We suppose that the connectivity (measured as directionless total flow) between a “home” unit *i* and a “hub” unit *j* during week *t* is given by the log-linear causal model:

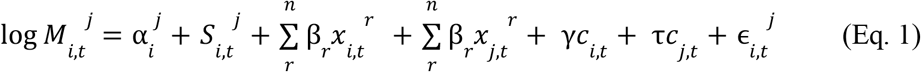

where a log-transformed dependent variable is used to allow the independent variables to influence connectivity 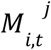 multiplicatively; also given that we are interested in the resulting difference in connectivity compared to some baseline value as a result of a change in case incidence, the relative change in connectivity has a more meaningful interpretation than an absolute one. 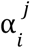 is a time-invariant measure of heterogeneity in connectivity between pairs of “home” and “hub” units; 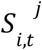 is a time trend that reflects potential seasonal changes in connectivity between “home” unit *i* and “hub” unit *j*; 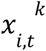 and 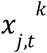 for *k* = {0, 1, …, *n*} represent each of the factors associated with “home” unit *i* and “hub” unit *j* respectively; *c*_*i,t*_ and *c*_*j,t*_ represent the weekly number of new COVID-19 cases per capita at “home” unit *i* and “hub” unit *j* respectively; 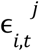 measures unobserved spatiotemporal factors that affect connectivity (modelled as a Gaussian).

(Eq. 1) motivates a difference-in-difference estimator (DID) for determining the causal effects of changes in COVID-19 case number per capita. We take the (first) time difference of Eq. 1, which results in the “temporal growth in connectivity” between “home” unit *i* and “hub” unit *j*; removing any time-constant individual home-hub effects. We then compute the difference in the temporal growth in connectivity between “home” unit *i* and “hub” unit *j* with the temporal growth between the same “home” unit *i* and a different “hub” unit *k*. This results in an equation of the form:

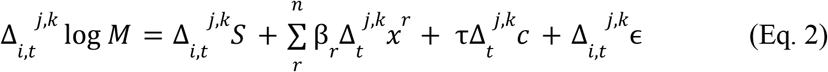

where 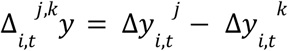, with 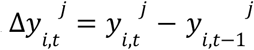, and 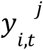 is a quantity associated with the connectivity between *i* and *j* observed in week *t*; 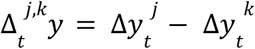, with 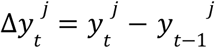 where 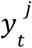 is a quantity associated with *j* observed in week *t*. The coefficient *τ* can be interpreted as the proportional change in the connectivity between *i* and *j* for a given week as a result of a unit increase in COVID-19 case number per capita at the “hub” *j*.

By aggregating the weekly mobility fluxes associated with each of the 30 “hub” units independently, we find that they share a broadly similar temporal profile characterised by a mostly stable period up until two weeks before Christmas when the level of mobility began to drop sharply across all “hubs” (Figure S9); this drop continued until the beginning of January 2022 when mobility began to recover over the course of the next few weeks. This observation motivates the assumption of homogeneity in the seasonal time trend in connectivity across different “hub” units for a given “home”, such that 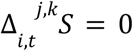, giving the final expression:

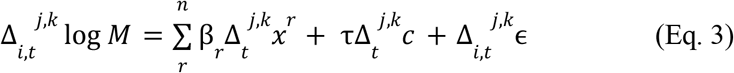

Future work should further assess the validity of this assumption considering mobility data from multiple years.

Note that with this triple-differences approach, we need only consider time-varying factors which vary across the “hub” units being considered, and any time-invariant factors (e.g. geographical distance and connectivity at baseline) and factors associated with the “home” unit can be ignored, so long as the functional form assumed in Eq. 1 is correct. These factors can however act as effect modifiers, as demonstrated in a subsequent model which takes into account the geographical distance between the “home” and “hub” unit (and similarly the proportion of population aged above 50 at the “home” unit), for which the model can be expressed as

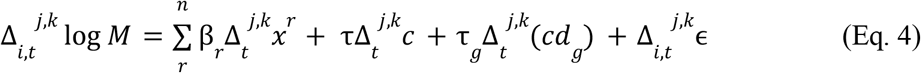

where 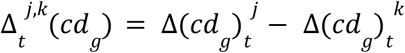, with 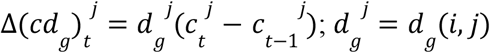 represents the geographical distance between the “home” unit *i* and “hub” unit *j*.

We fitted the above models using least-squares regression in R (version 4.0.2). To account for potential spatial correlation between “home” units that are in close proximity with each other, we computed the confidence intervals for the coefficient estimates using cluster-robust standard errors, with “home” units in the same Upper Tier Local Authority (UTLA) considered as clusters.

#### Bootstrapped confidence intervals and cluster analysis

The triple-differences approach of considering the difference in connectivity to different “hub” units for a given “home” unit creates redundant comparisons which may lead to overly confident standard errors. This can be illustrated using a simple example of a mobility network in which the “home” unit is connected to three “hub” units. Although there are three possible unique pairing of “hub” units, only two of them are independent with the remaining being a linear combination of the other two. To tackle this, following a similar approach as in Wymant el al. (*64*), we repeated the analysis with bootstrapping such that a subset of non-redundant comparison pairs were randomly sampled from all possible pairing for a given “home” unit. The confidence intervals for the coefficient estimates were then assessed from 5,000 bootstrap samples.

To again account for potential spatial correlation between “home” units, we further performed a cluster-bootstrap analysis in which “home” units that are in the same UTLA were considered as clusters. To create each bootstrap sample, we drew *n* of these UTLA clusters with replacement, where *n* is the number of “home” units without clustering. Within each UTLA, the same bootstrapping method as described above was applied to account for the additional uncertainty due to redundant comparison pairs. The confidence intervals for the coefficient estimates were then assessed from 5,000 bootstrap samples as before.

#### Sensitivity analyses

An important assumption made in this analysis is that connections to different “hub” units for a given “home” unit share a common seasonal time trend. We argue that this is a reasonable assumption during the Christmas holiday as most businesses close temporarily with the majority of the workers taking leaves (between only 2.1% - 4.0% and 3.3% - 6.0% of the labour force worked on Christmas Day and Boxing Day respectively across all regions in England, according to a census carried out by the Office of National Statistics in 2016 (https://www.ons.gov.uk/employmentandlabourmarket/peopleinwork/employmentandemployeetypes/adhocs/009371numberofpeopleworkingonchristmasdayandboxingdayuk2016)). The degree to which this assumption might be violated likely depends on the differences in the proportions of different employment sectors and types of businesses operating at the “hub” units, and therefore the nature of the observed mobility.

At the hub-level, we verified this assumption by considering the time trend in the aggregated mobility flux for each of the 30 “hub” units (Figure S9) and observed a broadly similar temporal profile across different “hub” units. We note however that Greater London appears to have suffered an earlier and more substantial drop in mobility compared to other “hub” units. This is perhaps not unexpected given the unique position of Greater London both in the mobility network and as a major business centre in England. Therefore as a sensitivity check, we repeated the above statistical analyses whilst ignoring all connections involving Greater London as a “hub” unit.

## Supplementary Information

**Figure S1:**
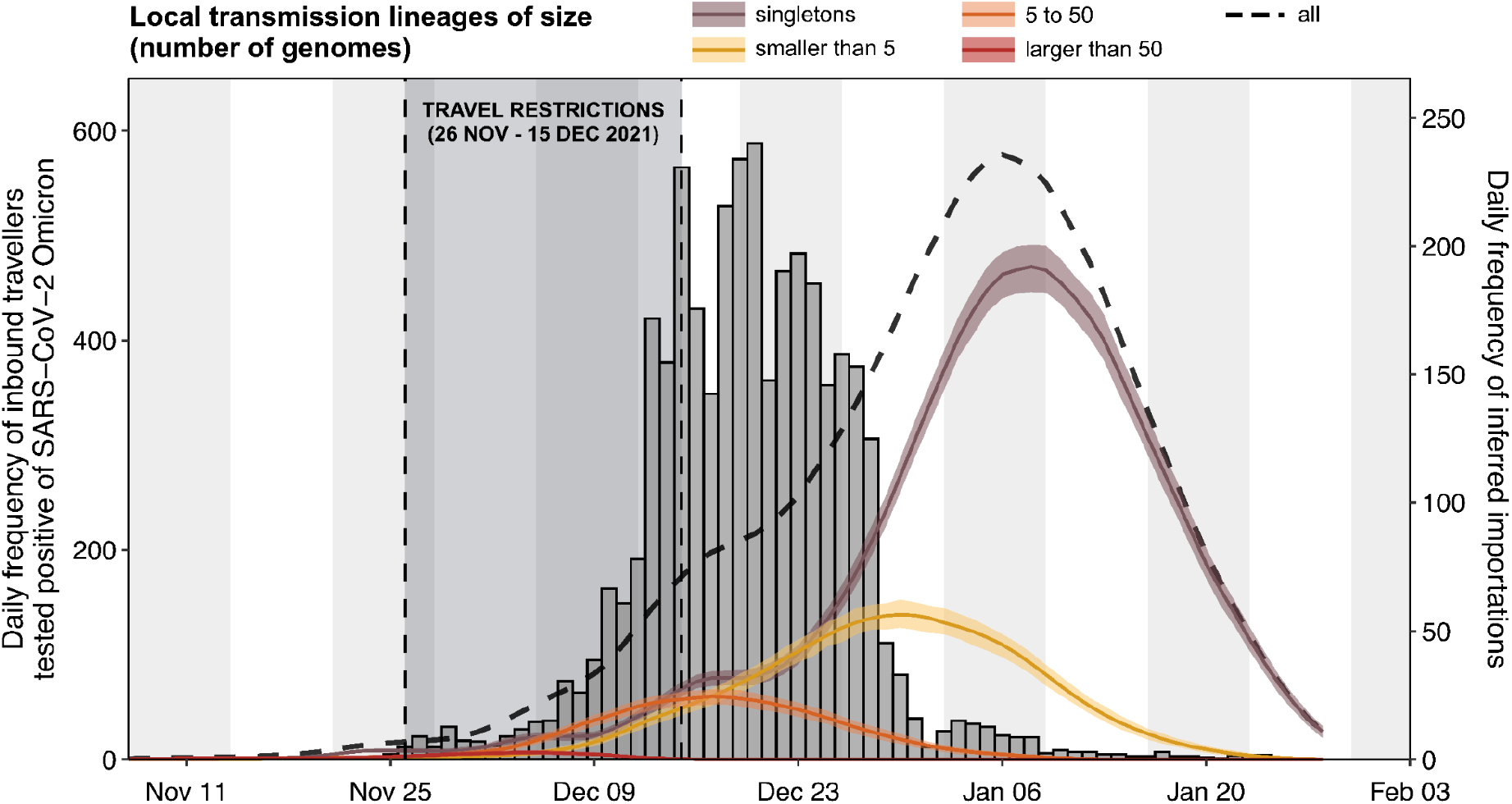
Comparison of Omicron BA.1 importation intensity as observed empirically from Variant Mutation (VAM) line list versus estimates from phylodynamic analysis. Related to Figure 1 and Figure 2. Histogram (grey bars) shows the daily number of incoming travellers who were later tested positive for Omicron BA.1 under the UKHSA mass testing programme (by specimen date). Solid lines represent the daily frequency of importations (7-day rolling average) as inferred from the phylodynamic analysis, coloured according to the size of resulting local transmission lineages (with the black dashed line representing the total numbers irrespective of size); shading denotes the 95% HPD across the posterior tree distribution.

**Figure S2.**
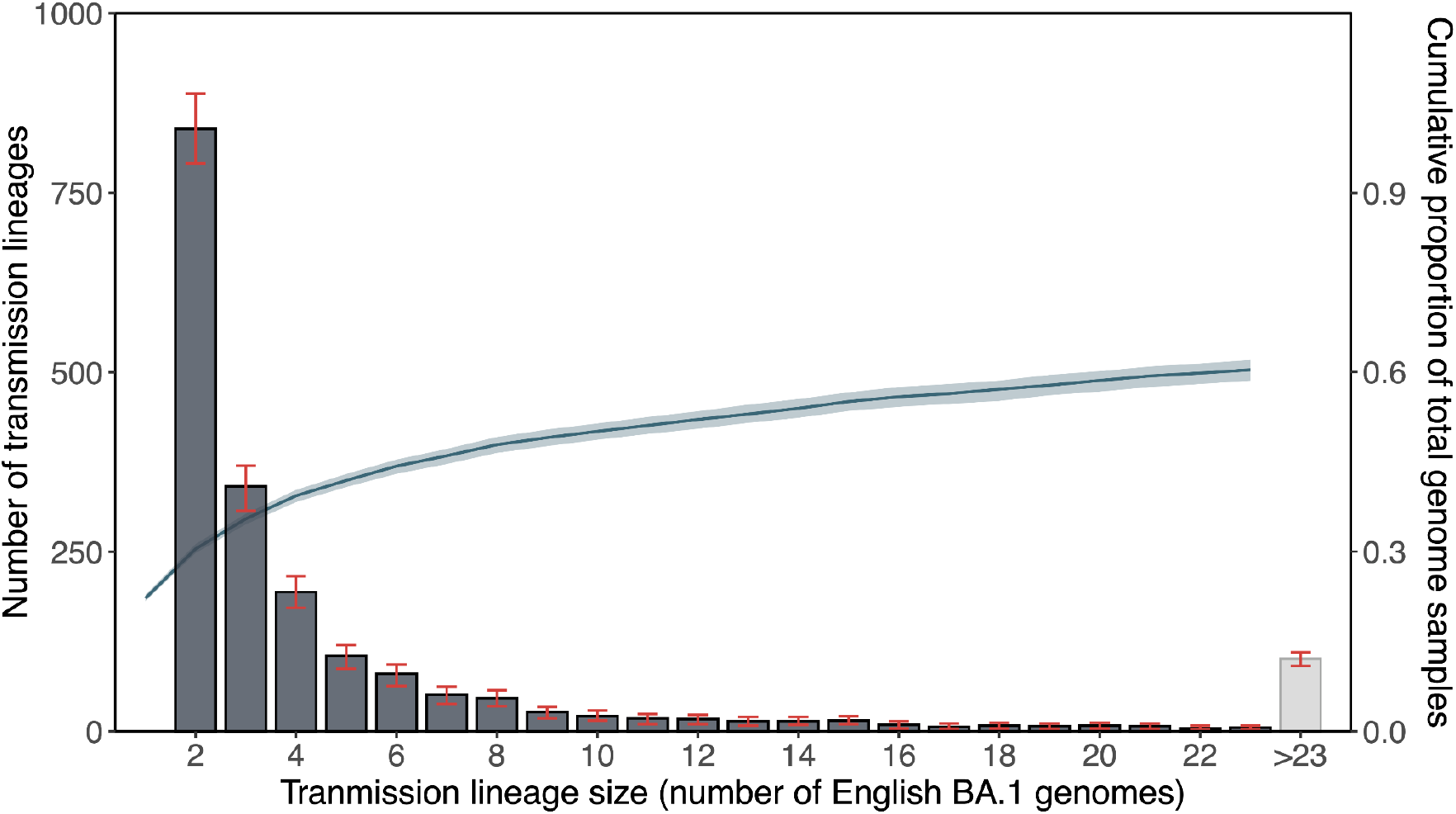
Distribution of local transmission lineage sizes from phylodynamic analysis. Related to Figure 1. Grey bars show the number of transmission lineages of different sizes; red error bars denote the 95% HPDs across the posterior tree distribution. Blue solid line represents the cumulative proportion of English Omicron BA.1 genomes in our dataset accounted for by transmission lineages up to a certain size; shading denotes the 95% HPD across the posterior tree distribution.

**Figure S3.**
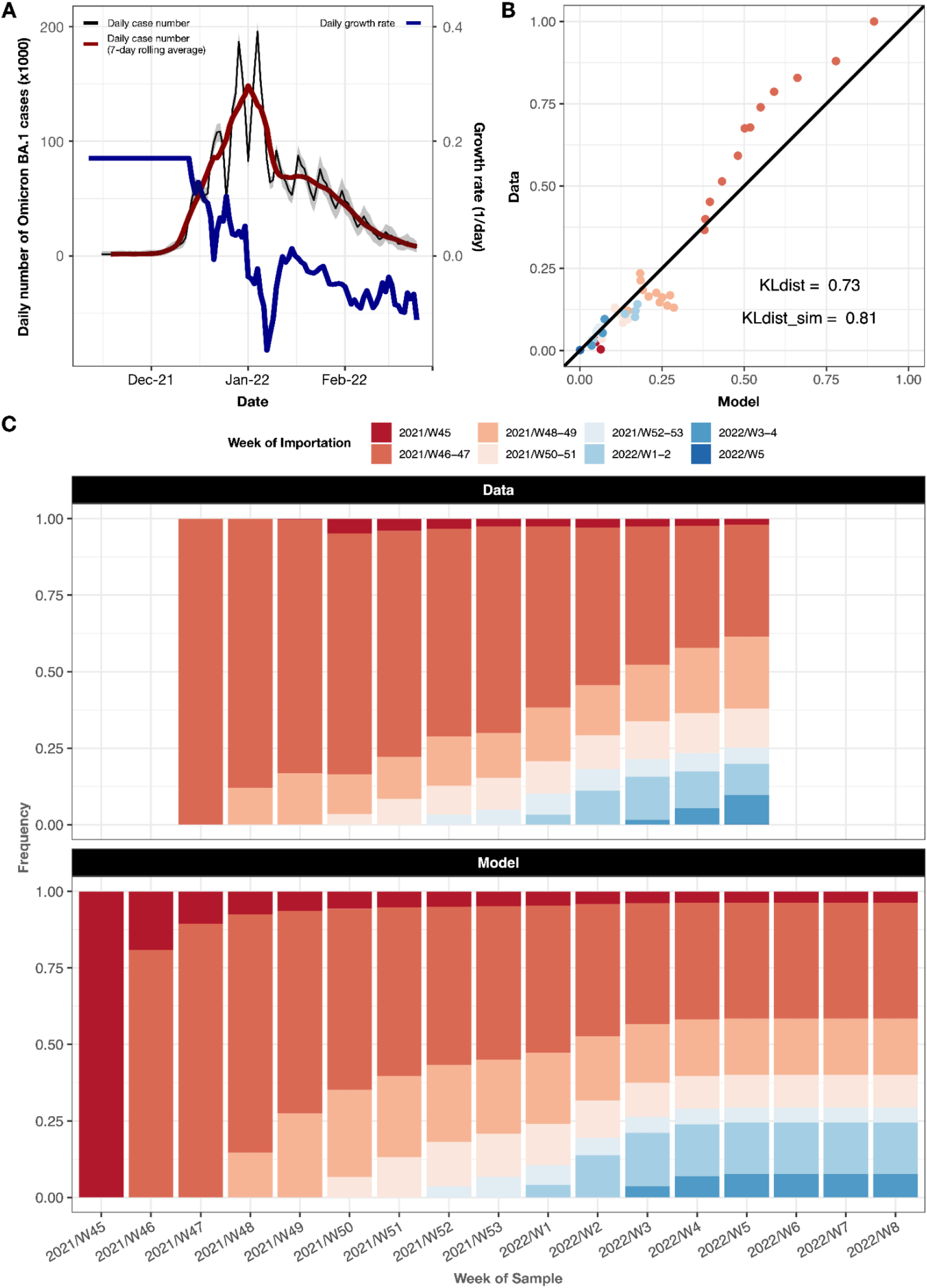
Comparison of transmission lineage size distribution from phylodynamic analysis versus simulated results from a branching process model. Related to Figure 1. (A) Black and red solid lines represent the estimated daily and 7-day rolling average daily number of Omicron BA.1 cases in England. Blue solid line represents the estimated daily growth rate, with the initial values imputed using an estimate of the growth rate on 13 December 2021. (B and C) Weekly proportion of local Omicron BA.1 infections resulting from importations at different times throughout the epidemic, with comparison between empirical observations from the phylodynamic analysis (C, top) and predictions from the branching process model (C, bottom).

**Figure S4.**
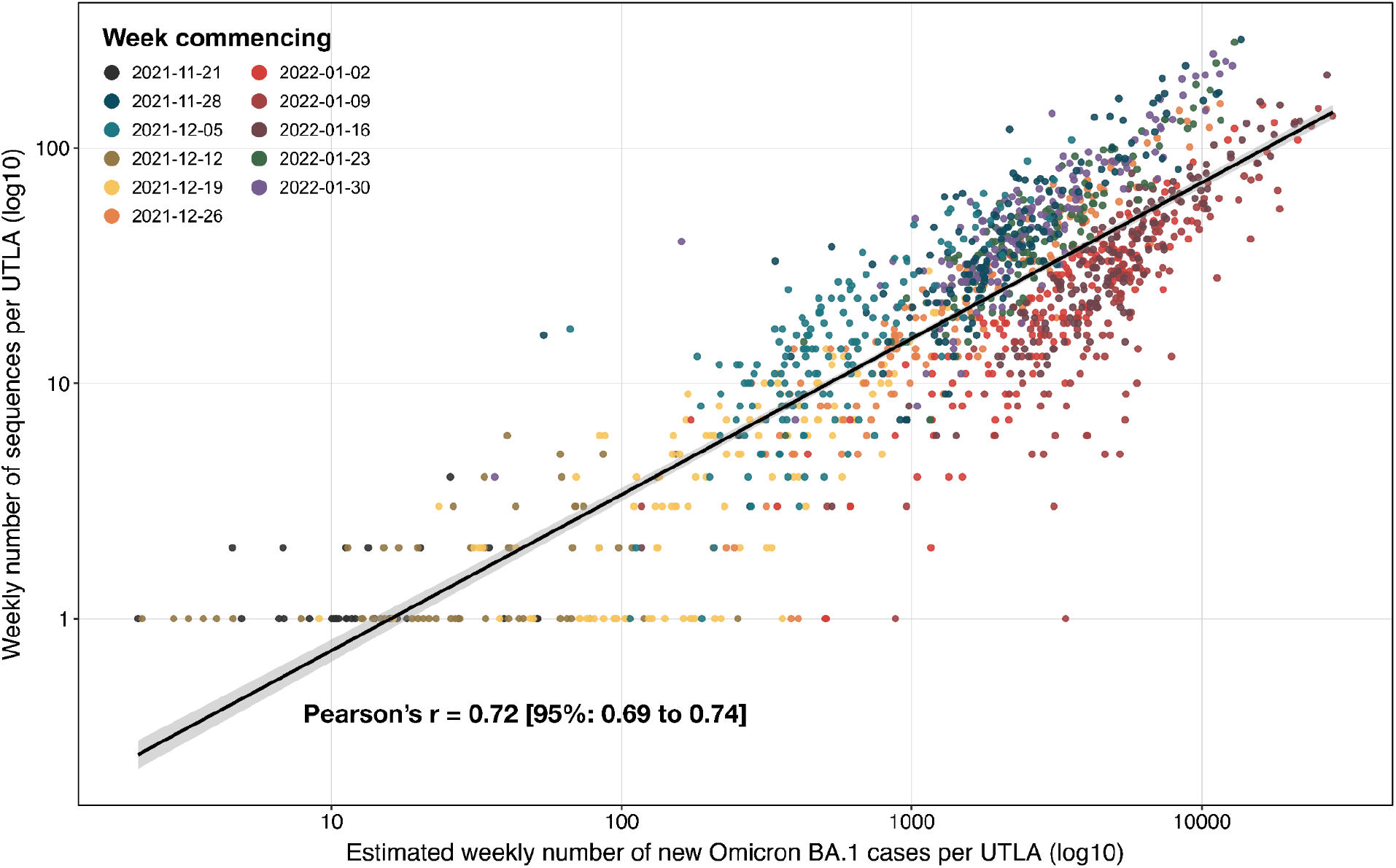
Correlation between estimated number of Omicron BA.1 cases and number of Omicron BA.1 genomes sampled across UTLAs in England. Related to Figure 3. Circles are coloured by week commencing date. Solid black line represents the line of best-fit; shaded region represents the 95% CI. We note in particular the clustering of circles corresponding to the same week along the line of best-fit, indicating small changes in sequencing coverage across time but not across UTLAs.

**Figure S5.**
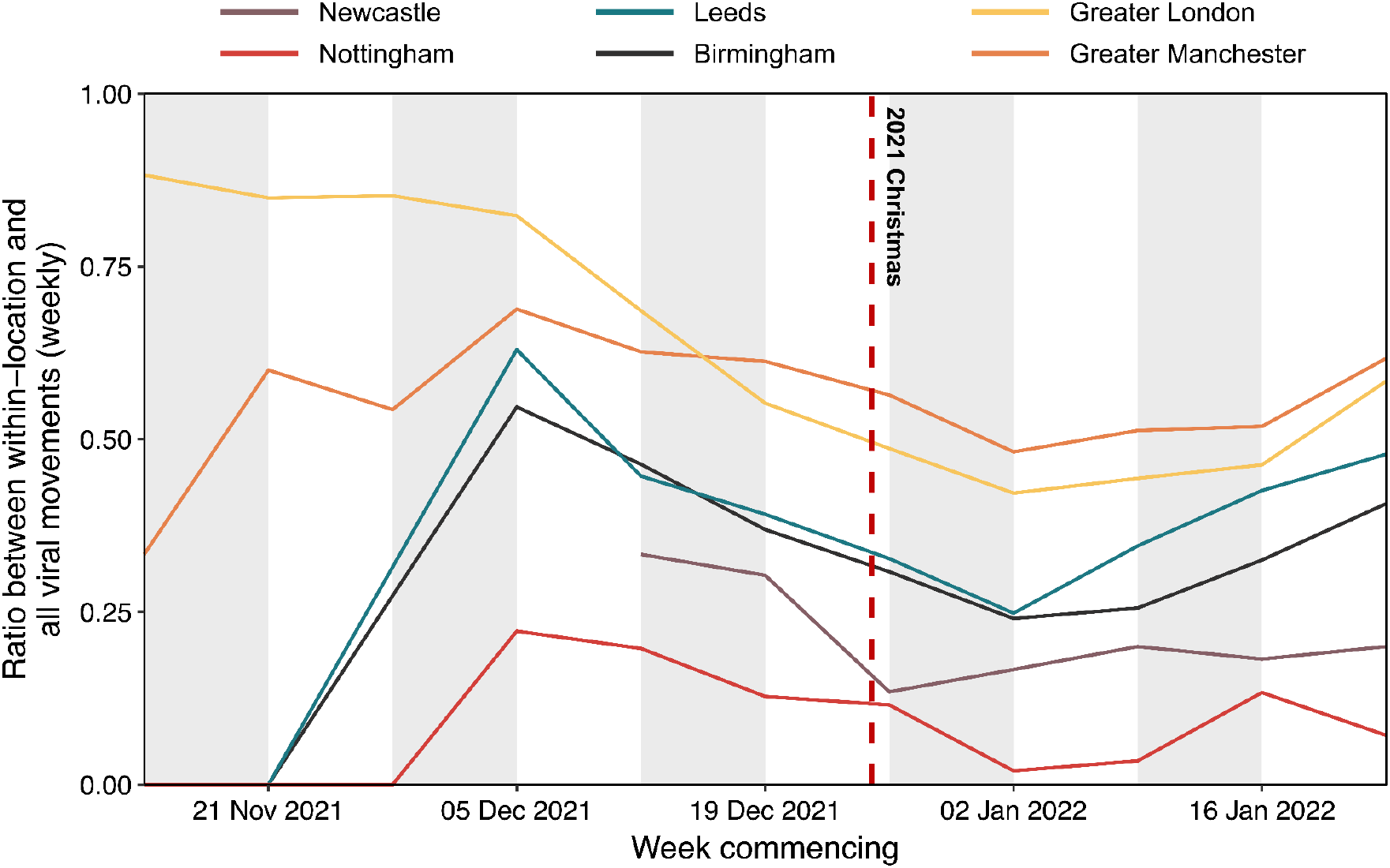
Within-location versus all viral lineage movements for major cities in England. Related to Figure 3 and 4. Each solid line represents the ratio between the frequency of within-location and all viral lineage movements per week, as inferred from continuous phylogeography for 6 major cities in England. For Greater Manchester and Greater London, viral lineages associated with multiple lower tier local authorities were aggregated in the calculation of these ratios. The timing of each viral lineage movement was assumed to be half-way between the inferred time of the nodes corresponding to the origin and destination.

**Figure S6.**
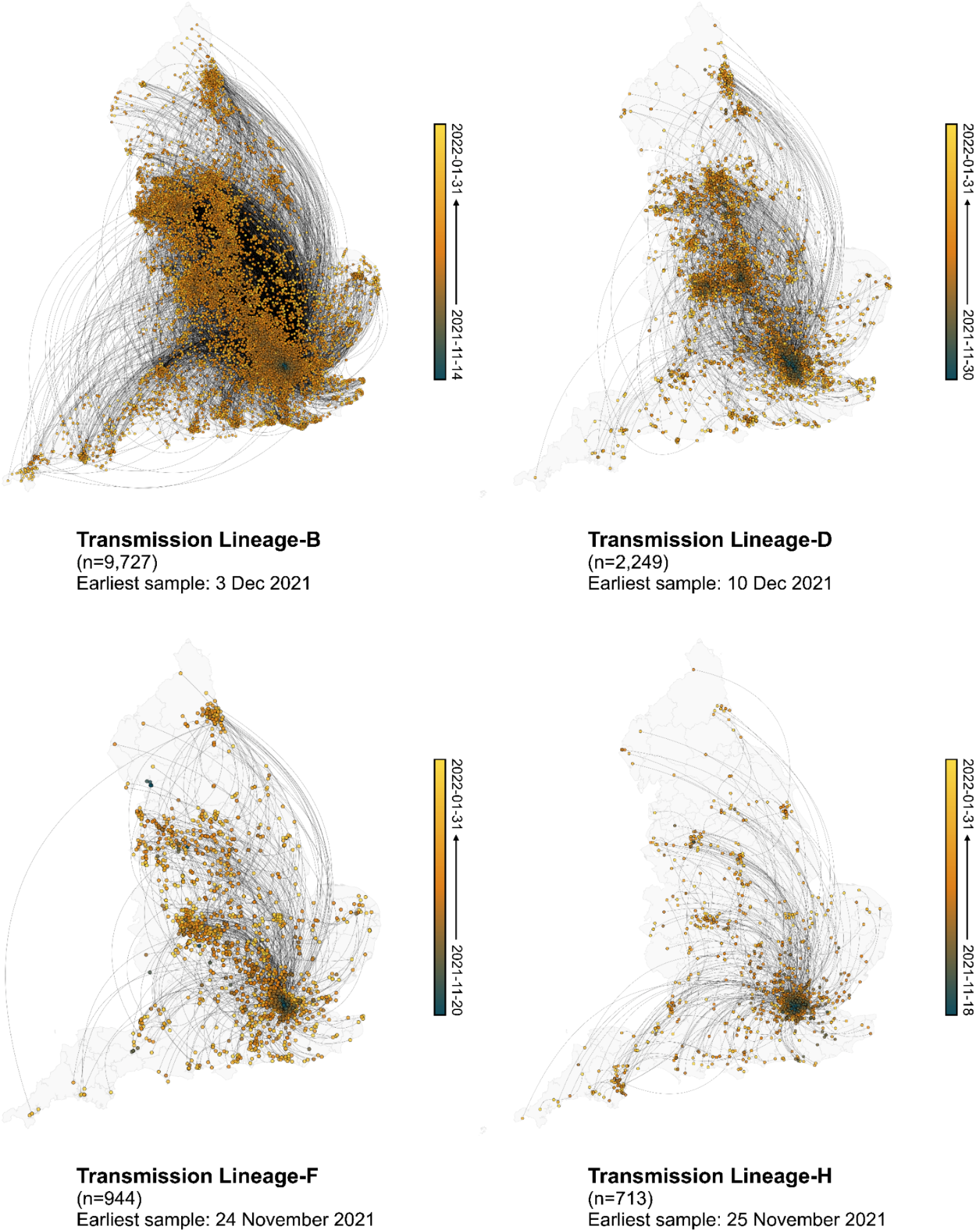
Spatiotemporal dynamics of Omicron BA.1 transmission lineages in England. Related to Figure 3. Maps showing viral lineage movements inferred from continuous phylogeography for Transmission Lineage-B, D, F and H. Nodes are coloured according to inferred date of occurrence and the direction of viral lineage movement is indicated by edge curvature (anti-clockwise).

**Figure S7.**
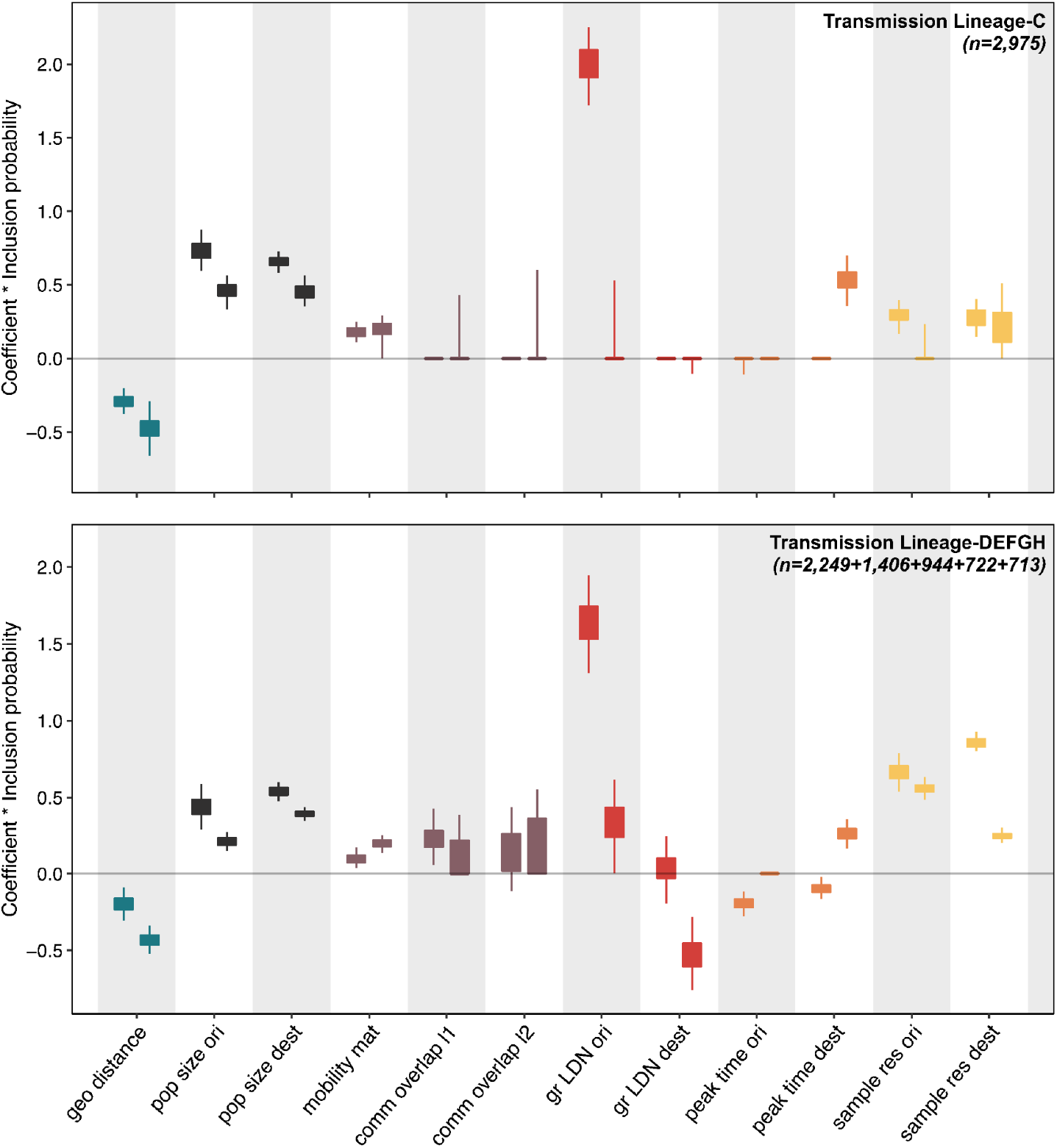
Predictors of viral lineage movements in England considered in the time-inhomogeneous discrete-phylogeography with GLM model. Related to Figure 4. Posterior distributions of the product of the log predictor coefficient and the predictor inclusion probability for each predictor. Estimates are plotted for Transmission Lineage-C (top) and Transmission Lineage-D, E, F, G and H (bottom) which were analysed together in a joint model. For each predictor, the left hand value represents the expansion phase, the right hand value the post-expansion phase. Posterior distributions are colored according to predictor type. Predictors include geographic distances (geo distance, red), population sizes at the origin and destination (pop size ori & pop size dest, yellow), aggregated mobility (mobility mat, dark blue), mobility-based community membership level 1 and level 2 (comm overlap l1 & l2, light blue), Greater London origin and destination (gr LDN ori & gr LDN dest, green), peak time at the origin and destination (peak time ori & peak time dest, yellow) and the residual of a regression of sample size against case count regression at either the origin and destination (sample res ori & sample res dest, grey).

**Figure S8.**
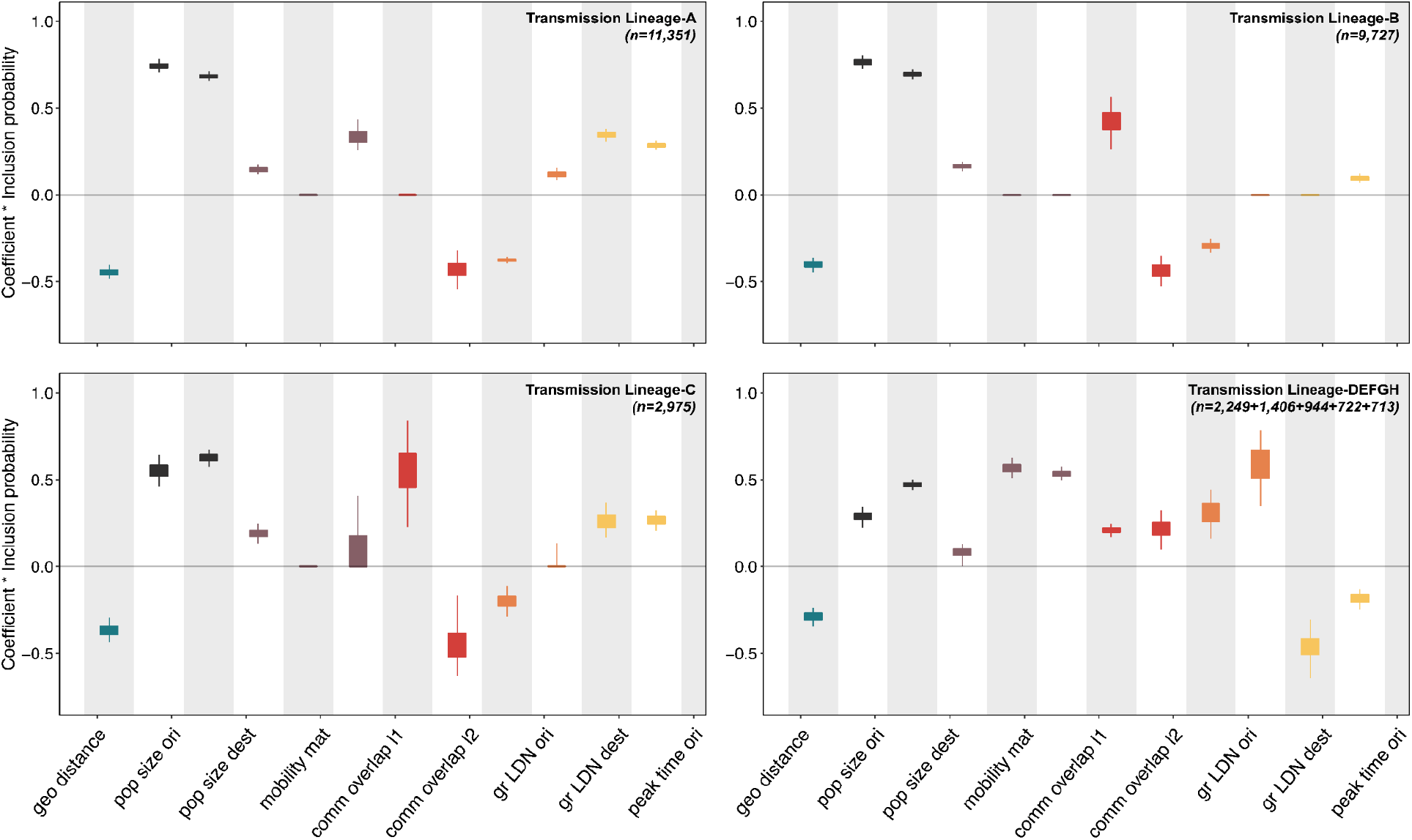
Predictors of viral lineage movements in England considered in the time-homogeneous discrete-phylogeography with GLM model. Related to Figure 4. Posterior distributions of the product of the log predictor coefficient and the predictor inclusion probability for each predictor. Estimates for the 8 largest transmission lineages are plotted, with the bottom-right panel showing estimates for Transmission Lineage-D, E, F, G and H estimated from a joint model. Posterior distributions are colored according to predictor type. Predictors include geographic distances (geo distance, red), population sizes at the origin and destination (pop size ori & pop size dest, yellow), aggregated mobility (mobility mat, dark blue), mobility-based community membership level 1 and level 2 (comm overlap l1 & l2, light blue), Greater London origin and destination (gr LDN ori & gr LDN dest, green), peak time at the origin and destination (peak time ori & peak time dest, yellow) and the residual of a regression of sample size against case count regression at either the origin and destination (sample res ori & sample res dest, grey).

**Figure S9.**
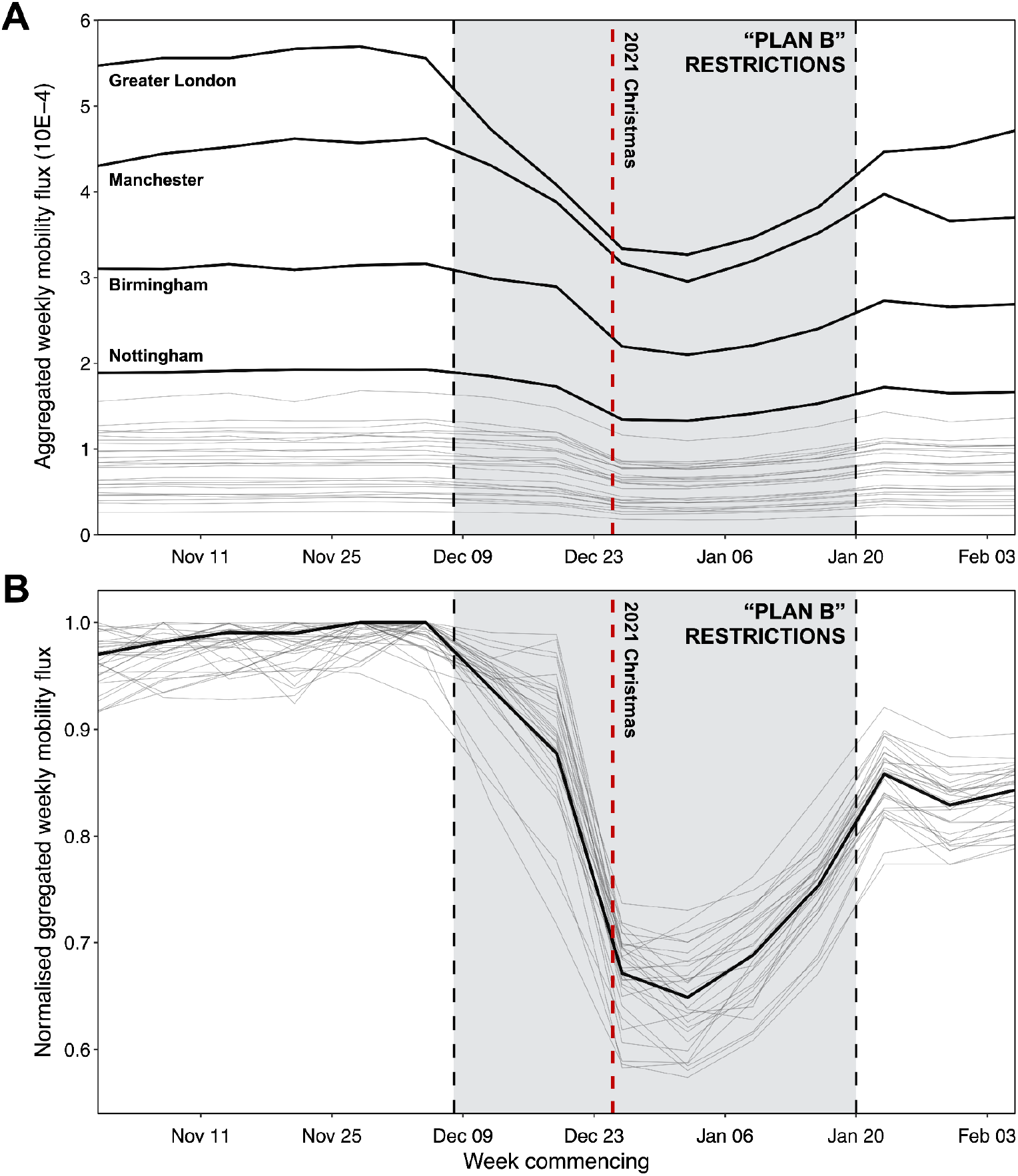
Mobility patterns associated with “hub” units. Related to Figure 5. (A) Weekly relative mobility to and from each of the 30 geographical units identified as “hubs” in the statistical analysis. (B) Weekly relative mobility to and from each of the 30 geographical units classified as “hubs”, normalised by the maximum observed value between November 2021 and January 2022.

**Table S1.** Imputation of initial growth rate in branch process model. Kullback-Leibler (KL) divergence values computed from comparisons of the weekly proportion of local Omicron BA.1 infections resulting from importations at different times as inferred from the phylodynamic analysis, versus predictions from branching process models with a range of initial growth rates. The initial growth rates in the branching models were imputed using estimates taken from the early growth phase of the epidemic (between 2021-12-04 and 2021-12-15), and the growth rate that minimised the KL divergence (shown in bold) was used in the best-fit model.

| Initial growth rate taken from | Kullback–Leibler divergence |
| --- | --- |
| 2021-12-04 | 2.84 |
| 2021-12-05 | 2.04 |
| 2021-12-06 | 1.33 |
| 2021-12-07 | 1.42 |
| 2021-12-08 | 2.95 |
| 2021-12-09 | 12.54 |
| 2021-12-10 | 13.13 |
| 2021-12-11 | 8.31 |
| 2021-12-12 | 2.02 |
| 2021-12-13 | **0.81** |
| 2021-12-14 | 2.76 |
| 2021-12-15 | 5.17 |

**Table S2.** Predictors of viral lineage movements in England. Related to Figure 4. Summary table and descriptions of predictors considered in the discrete phylogeographic GLM analysis. For location-specific predictors (as indicated by an asterisk), we included both an origin and a destination covariate in the GLM model.

| Predictor | Domain | Time-varying | Description |
| --- | --- | --- | --- |
| **Geographical distance between origin and destination** | Geography | No | Geographical distance between the origin and destination calculated using the Haversine formula in km |
| **Population size\*** | Demography | No | Population size at the origin/destination, from population estimates obtained by the Office of National Statistics in mid-year 2020 |
| **Aggregated mobility matrix** | Human mobility | Yes | Average weekly number of trips taken between origin and destination estimated from Google COVID-19 Aggregated Mobility Research Dataset; given that the mobility flux between two locations does not in general differ from symmetry in a statistically significant manner (i.e. the magnitude of mobility flux in either direction is generally very similar for a given connection), we considered a mobility matrix that was symmetrised |
| **Community memberships from mobility network (level-1/2)\*** | Human mobility | Yes | A binary variable [0,1] indicating whether the origin and destination belong to the same community at level-1/2; community structures were identified from the human mobility network as described by the aggregated mobility matrix, using the community detection algorithm Infomap (65, 66), with level-1/2 corresponding to the tree-depth at which the communities were extracted; level-1 has a higher level of aggregation (fewer communities) compared to level-2 (more communities) |
| **Greater London / non-Greater London indicator\*** | Geography/<br>Epidemiology | No | A binary variable [0,1] indicating whether the origin/destination is in the Greater London region |
| **Timing of peak in Omicron BA.1 case incidence\*** | Epidemiology | No | Timing of first peak in Omicron BA.1 case incidence at the origin/destination, measured as the number of days from 1 December 2021 |
| **Sampling residuals\*** | Sampling | No | Residuals from a regression of sample size against Omicron BA.1 case count at origin/destination; time-invariant residuals were used due to the small number of samples in some locations during the post-expansion phase |

**Table S3.** Constituent lower tier local authorities (LTLAs) of “home” and “hub” units identified in statistical analysis. Related to Figure 5. Table of the 30 and 105 geographical units that were labelled as “home” and “hub” units respectively (with the study period defined as between the week commencing on 28 November 2021 and the week ending on 2 January 2022), and their constituent LTLAs. Names of the geographical units are noted where multiple LTLAs have been aggregated.

| Unit type | Aggregation | Lower tier local authorities (LTLAs) |
| --- | --- | --- |
| Hub (n=30) | Greater London (n=32) | E09000002, E09000016, E09000026, E09000025, E09000030, E09000028, E09000006, E09000004, E09000011, E09000023, E09000008, E09000022, E09000024, E09000021, E09000027, E09000013, E09000005, E09000003, E09000007, E09000014, E09000010, E09000031, E09000012, E09000019, E09000033, E09000020, E09000032, E09000015, E09000009, E09000017, E09000018, E09000029 |
|  | Bristol and South Gloucestershire (n=2) | E06000023, E06000025 |
|  | Leeds and Bradford (n=2) | E08000032, E08000035 |
|  | Leicester and Blaby (n=2) | E06000016, E07000129 |
|  | Liverpool and Knowsley (n=2) | E08000011, E08000012 |
|  | N/A (n=25) | E06000002, E06000010, E06000015, E06000018, E06000021, E06000026, E06000030, E06000032, E06000033, E06000038, E06000042, E06000043, E06000044, E06000045, E06000058, E07000148, E07000154, E08000001, E08000003, E08000019, E08000021, E08000024, E08000025, E08000026, E08000031 |
| Home (study period defined as 28 November 2021 to 2 January 2022, inclusive) (n=105) | N/A (n=105) | E06000005, E06000007, E06000014, E06000020, E06000022, E06000031, E06000034, E06000036, E06000037, E06000039, E06000040, E06000041, E06000047, E06000049, E06000050, E06000051, E06000054, E06000055, E06000056, E06000060, E07000008, E07000009, E07000033, E07000034, E07000036, E07000041, E07000045, E07000061, E07000063, E07000066, E07000069, E07000070, E07000071, E07000075, E07000078, E07000081, E07000084, E07000085, E07000086, E07000087, E07000088, E07000089, E07000090, E07000091, E07000093, E07000094, E07000096, E07000099, E07000103, E07000118, E07000121, E07000123, E07000130, E07000132, E07000134, E07000138, E07000141, E07000150, E07000151, E07000153, E07000155, E07000156, E07000164, E07000171, E07000172, E07000175, E07000177, E07000178, E07000179, E07000192, E07000193, E07000194, E07000197, E07000199, E07000202, E07000209, E07000211, E07000213, E07000218, E07000219, E07000220, E07000222, E07000226, E07000228, E07000229, E07000237, E07000240, E07000241, E07000243, E07000246, E08000002, E08000006, E08000007, E08000009, E08000010, E08000016, E08000017, E08000022, E08000027, E08000028, E08000029, E08000030, E08000034, E08000036, E08000037 |

**Table S4.**
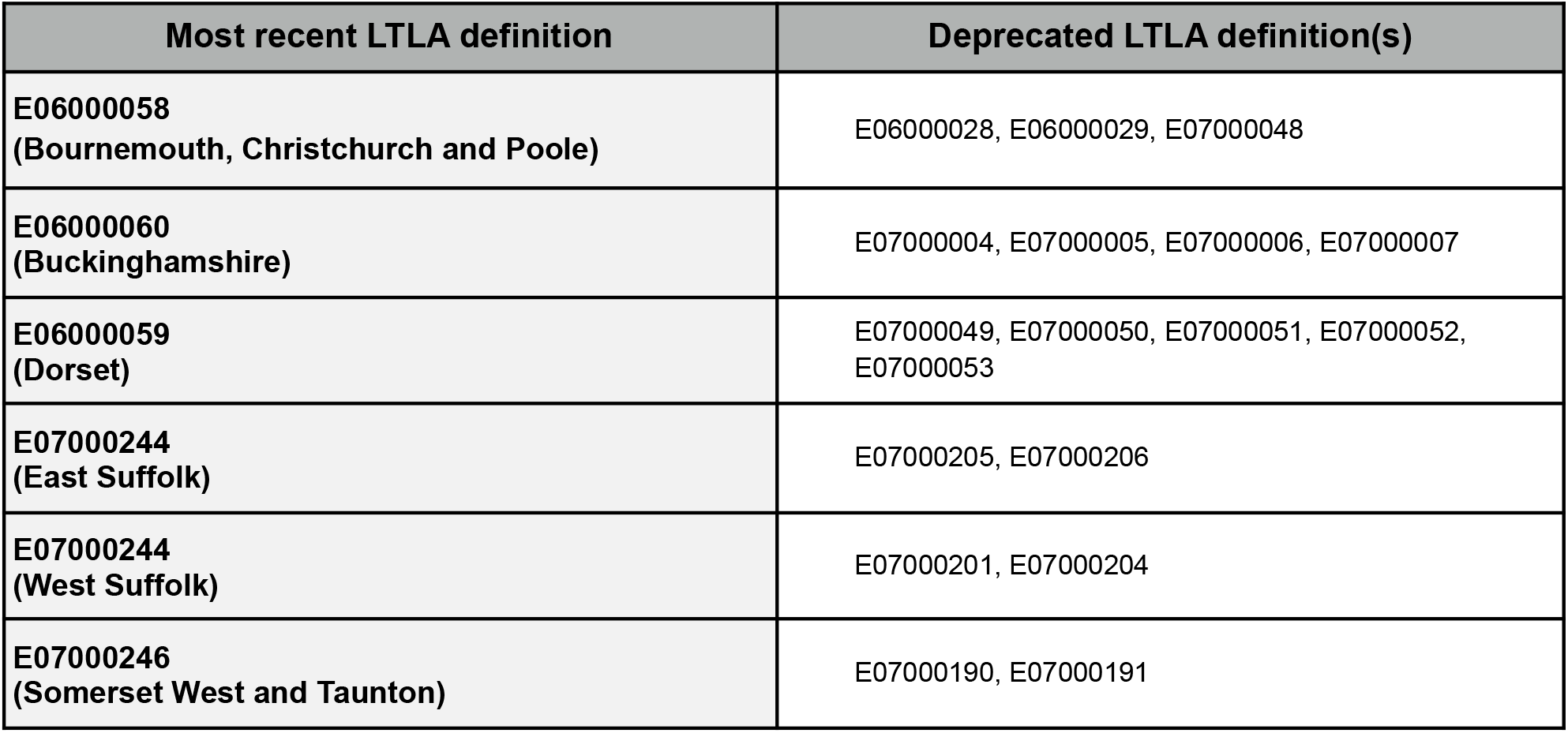
Definitions of lower tier local authorities. Table of LTLAs that have been deprecated and aggregated into newly defined LTLAs, according to recent definitions used in the report of population estimates for the UK in mid-2020, compiled by the Office of National Statistics, UK.

| Most recent LTLA definition | Deprecated LTLA definition(s) |
| --- | --- |
| **E06000058**<br>(Bournemouth, Christchurch and Poole) | E06000028, E06000029, E07000048 |
| **E06000060**<br>(Buckinghamshire) | E07000004, E07000005, E07000006, E07000007 |
| **E06000059**<br>(Dorset) | E07000049, E07000050, E07000051, E07000052, E07000053 |
| **E07000244**<br>(East Suffolk) | E07000205, E07000206 |
| **E07000244**<br>(West Suffolk) | E07000201, E07000204 |
| **E07000246**<br>(Somerset West and Taunton) | E07000190, E07000191 |

**Table S5.** Results from statistical model estimating the impact of changes in COVID-19 case incidence on connectivity to “hub” units. The effect is measured as the percent reduction in connectivity (total mobility flow regardless of direction) between a “home” and a “hub” unit, for every unit increase in COVID-19 case number per 100 people at the “hub”. Numbers in bold are coefficient estimates from least-squares regression; ranges (in brackets) shown next to the coefficient estimates are 95% confidence intervals (CIs) from least-squares regression without accounting for spatial correlation between “home” units; ranges shown below the coefficient estimates (in the same row) are 95% CIs with cluster-robust standard errors at the upper tier local authority (UTLA) level. 95% CIs assessed from bootstrap analysis (5,000 bootstrap samples) with (bottom) and without (top) considering spatial clustering of “home” units at the UTLA level are also shown for each coefficient estimate. The study period is defined as the period from the week commencing on 28 November 2021 to the week ending on 1 January 2022.

| Percent reduction in connectivity between a “home” unit and a “hub” unit for every unit increase in non-variant-specific COVID-19 case number per 100 people at the “hub” |  |
| --- | --- |
| All “hubs” units included | **12.14** (8.14 to 15.96)<br>(with UTLA-CRSE: 5.22 to 18.55) |
| Bootstrap analysis | (7.66 to 16.54)<br>(UTLA-clustered: 2.84 to 20.81) |
| Greater London excluded | **17.40** (7.43 to 26.30)<br>(with UTLA-CRSE: 0.14 to 31.67) |
| Bootstrap analysis | (6.56 to 27.08)<br>(UTLA-clustered: -5.66 to 37.22) |
| Percent reduction in connectivity between a “home” unit and a “hub” unit for every unit increase in non-variant-specific COVID-19 case number per 100 people at the “hub”, with geographical distance between “home” and “hub” as effect modifier |  |
| All “hubs” units included<br>(geographical distance of 0 km) | **24.04** (18.98 to 28.78)<br>(with UTLA-CRSE: 17.99 to 29.65) |
| Bootstrap analysis | (18.61 to 29.21)<br>(UTLA-clustered: 14.43 to 32.93) |
| All “hubs” units included<br>(geographical distance of 50 km) | **20.02** (15.63 to 24.18)<br>(with UTLA-CRSE: 14.50 to 25.18) |
| Bootstrap analysis | (15.16 to 24.79)<br>(UTLA-clustered: 11.49 to 28.36) |
| All “hubs” units included<br>(geographical distance of 100 km) | **15.79** (11.81 to 19.58)<br>(with UTLA-CRSE: 10.29 to 20.94) |
| Bootstrap analysis | (11.29 to 20.17)<br>(UTLA-clustered: 7.70 to 23.94) |
| Greater London excluded<br>(geographical distance of 0 km) | **23.32** (12.67 to 32.68)<br>(with UTLA-CRSE: 5.68 to 37.67) |
| Bootstrap analysis | (12.59 to 32.80)<br>(UTLA-clustered: -2.11 to 42.62) |
| Greater London excluded<br>(geographical distance of 50 km) | **16.72** (6.68 to 25.67)<br>(with UTLA-CRSE: -0.04 to 30.67) |
| Bootstrap analysis | (6.38 to 25.89)<br>(UTLA-clustered: -4.09 to 36.89) |
| **Greater London excluded**<br>**(geographical distance of 100 km)** | **9.54** (-3.78 to 21.15)<br>(with UTLA-CRSE: -11.58 to 26.66) |
| **Bootstrap analysis** | (-0.02 to 20.08)<br>(UTLA-clustered: -14.32 to 34.86) |

***Table S6. The COVID-19 Genomics UK (COG-UK) consortium, June 2021 V4***

**The COVID-19 Genomics UK (COG-UK) consortium FINAL (06-2021 V4)**

**Funding acquisition, Leadership and supervision, Metadata curation, Project administration, Samples and logistics, Sequencing and analysis, Software and analysis tools, and Visualisation:**

Samuel C Robson ^13, 84^

**Funding acquisition, Leadership and supervision, Metadata curation, Project administration, Samples and logistics, Sequencing and analysis, and Software and analysis tools:**

Thomas R Connor ^11, 74^ and Nicholas J Loman ^43^

**Leadership and supervision, Metadata curation, Project administration, Samples and logistics, Sequencing and analysis, Software and analysis tools, and Visualisation:**

Tanya Golubchik ^5^

**Funding acquisition, Leadership and supervision, Metadata curation, Samples and logistics, Sequencing and analysis, and Visualisation:**

Rocio T Martinez Nunez ^46^

**Funding acquisition, Leadership and supervision, Project administration, Samples and logistics, Sequencing and analysis, and Software and analysis tools:**

David Bonsall ^5^

**Funding acquisition, Leadership and supervision, Project administration, Sequencing and analysis, Software and analysis tools, and Visualisation:**

Andrew Rambaut ^104^

**Funding acquisition, Metadata curation, Project administration, Samples and logistics, Sequencing and analysis, and Software and analysis tools:**

Luke B Snell ^12^

**Leadership and supervision, Metadata curation, Project administration, Samples and logistics, Software and analysis tools, and Visualisation:**

Rich Livett ^116^

**Funding acquisition, Leadership and supervision, Metadata curation, Project administration, and Samples and logistics:**

Catherine Ludden ^20, 70^

**Funding acquisition, Leadership and supervision, Metadata curation, Samples and logistics, and Sequencing and analysis:**

Sally Corden ^74^ and Eleni Nastouli ^96, 95, 30^

**Funding acquisition, Leadership and supervision, Metadata curation, Sequencing and analysis, and Software and analysis tools:**

Gaia Nebbia ^12^

**Funding acquisition, Leadership and supervision, Project administration, Samples and logistics, and Sequencing and analysis:**

Ian Johnston ^116^

**Leadership and supervision, Metadata curation, Project administration, Samples and logistics, and Sequencing and analysis:**

Katrina Lythgoe ^5^, M. Estee Torok ^19, 20^ and Ian G Goodfellow ^24^

**Leadership and supervision, Metadata curation, Project administration, Samples and logistics, and Visualisation:**

Jacqui A Prieto ^97, 82^ and Kordo Saeed ^97, 83^

**Leadership and supervision, Metadata curation, Project administration, Sequencing and analysis, and Software and analysis tools:**

David K Jackson ^116^

**Leadership and supervision, Metadata curation, Samples and logistics, Sequencing and analysis, and Visualisation:**

Catherine Houlihan ^96, 94^

**Leadership and supervision, Metadata curation, Sequencing and analysis, Software and analysis tools, and Visualisation:**

Dan Frampton ^94, 95^

**Metadata curation, Project administration, Samples and logistics, Sequencing and analysis, and Software and analysis tools:**

William L Hamilton ^19^ and Adam A Witney ^41^

**Funding acquisition, Samples and logistics, Sequencing and analysis, and Visualisation:**

Giselda Bucca ^101^

**Funding acquisition, Leadership and supervision, Metadata curation, and Project administration:**

Cassie F Pope ^40, 41^

**Funding acquisition, Leadership and supervision, Metadata curation, and Samples and logistics:**

Catherine Moore ^74^

**Funding acquisition, Leadership and supervision, Metadata curation, and Sequencing and analysis:**

Emma C Thomson ^53^

**Funding acquisition, Leadership and supervision, Project administration, and Samples and logistics:**

Teresa Cutino-Moguel ^2^, Ewan M Harrison ^116, 102^

**Funding acquisition, Leadership and supervision, Sequencing and analysis, and Visualisation:**

Colin P Smith ^101^

**Leadership and supervision, Metadata curation, Project administration, and Sequencing and analysis:**

Fiona Rogan ^77^

**Leadership and supervision, Metadata curation, Project administration, and Samples and logistics:**

Shaun M Beckwith ^6^, Abigail Murray ^6^, Dawn Singleton ^6^, Kirstine Eastick ^37^, Liz A Sheridan ^98^, Paul Randell ^99^, Leigh M Jackson ^105^, Cristina V Ariani ^116^ and Sónia Gonçalves ^116^

**Leadership and supervision, Metadata curation, Samples and logistics, and Sequencing and analysis:**

Derek J Fairley ^3, 77^, Matthew W Loose ^18^ and Joanne Watkins ^74^

**Leadership and supervision, Metadata curation, Samples and logistics, and Visualisation:**

Samuel Moses ^25, 106^

**Leadership and supervision, Metadata curation, Sequencing and analysis, and Software and analysis tools:**

Sam Nicholls ^43^, Matthew Bull ^74^ and Roberto Amato ^116^

**Leadership and supervision, Project administration, Samples and logistics, and Sequencing and analysis:**

Darren L Smith ^36, 65, 66^

**Leadership and supervision, Sequencing and analysis, Software and analysis tools, and Visualisation:**

David M Aanensen ^14, 116^ and Jeffrey C Barrett ^116^

**Metadata curation, Project administration, Samples and logistics, and Sequencing and analysis:**

Beatrix Kele ^2^, Dinesh Aggarwal ^20, 116, 70^, James G Shepherd ^53^, Martin D Curran ^71^ and Surendra Parmar ^71^

**Metadata curation, Project administration, Sequencing and analysis, and Software and analysis tools:**

Matthew D Parker ^109^

**Metadata curation, Samples and logistics, Sequencing and analysis, and Software and analysis tools:**

Catryn Williams ^74^

**Metadata curation, Samples and logistics, Sequencing and analysis, and Visualisation:**

Sharon Glaysher ^68^

**Metadata curation, Sequencing and analysis, Software and analysis tools, and Visualisation:**

Anthony P Underwood ^14, 116^, Matthew Bashton ^36, 65^, Nicole Pacchiarini ^74^, Katie F Loveson ^84^ and Matthew Byott ^95, 96^

**Project administration, Sequencing and analysis, Software and analysis tools, and Visualisation:**

Alessandro M Carabelli ^20^

**Funding acquisition, Leadership and supervision, and Metadata curation:**

Kate E Templeton ^56, 104^

**Funding acquisition, Leadership and supervision, and Project administration:**

Sharon J Peacock ^20, 70^, Thushan I de Silva ^109^, Dennis Wang ^109^, Cordelia F Langford ^116^ and John Sillitoe ^116^

**Funding acquisition, Leadership and supervision, and Samples and logistics:**

Rory N Gunson ^55^

**Funding acquisition, Leadership and supervision, and Sequencing and analysis:**

Simon Cottrell ^74^, Justin O’Grady ^75, 103^ and Dominic Kwiatkowski ^116, 108^

**Leadership and supervision, Metadata curation, and Project administration:**

Patrick J Lillie ^37^

**Leadership and supervision, Metadata curation, and Samples and logistics:**

Nicholas Cortes ^33^, Nathan Moore ^33^, Claire Thomas ^33^, Phillipa J Burns ^37^, Tabitha W Mahungu ^80^ and Steven Liggett ^86^

**Leadership and supervision, Metadata curation, and Sequencing and analysis:**

Angela H Beckett ^13, 81^ and Matthew TG Holden ^73^

**Leadership and supervision, Project administration, and Samples and logistics:**

Lisa J Levett ^34^, Husam Osman ^70, 35^ and Mohammed O Hassan-Ibrahim ^99^

**Leadership and supervision, Project administration, and Sequencing and analysis:**

David A Simpson ^77^

**Leadership and supervision, Samples and logistics, and Sequencing and analysis:**

Meera Chand ^72^, Ravi K Gupta ^102^, Alistair C Darby ^107^ and Steve Paterson ^107^

**Leadership and supervision, Sequencing and analysis, and Software and analysis tools:**

Oliver G Pybus ^23^, Erik M Volz ^39^, Daniela de Angelis ^52^, David L Robertson ^53^, Andrew J Page ^75^ and Inigo Martincorena ^116^

**Leadership and supervision, Sequencing and analysis, and Visualisation:**

Louise Aigrain ^116^ and Andrew R Bassett ^116^

**Metadata curation, Project administration, and Samples and logistics:**

Nick Wong ^50^, Yusri Taha ^89^, Michelle J Erkiert ^99^ and Michael H Spencer Chapman ^116, 102^

**Metadata curation, Project administration, and Sequencing and analysis:**

Rebecca Dewar ^56^ and Martin P McHugh ^56, 111^

**Metadata curation, Project administration, and Software and analysis tools:**

Siddharth Mookerjee ^38, 57^

**Metadata curation, Project administration, and Visualisation:**

Stephen Aplin ^97^, Matthew Harvey ^97^, Thea Sass ^97^, Helen Umpleby ^97^ and Helen Wheeler ^97^

**Metadata curation, Samples and logistics, and Sequencing and analysis:**

James P McKenna ^3^, Ben Warne ^9^, Joshua F Taylor ^22^, Yasmin Chaudhry ^24^, Rhys Izuagbe ^24^, Aminu S Jahun ^24^, Gregory R Young ^36, 65^, Claire McMurray ^43^, Clare M McCann ^65, 66^, Andrew Nelson ^65, 66^ and Scott Elliott ^68^

**Metadata curation, Samples and logistics, and Visualisation:**

Hannah Lowe ^25^

**Metadata curation, Sequencing and analysis, and Software and analysis tools:**

Anna Price ^11^, Matthew R Crown ^65^, Sara Rey ^74^, Sunando Roy ^96^ and Ben Temperton ^105^

**Metadata curation, Sequencing and analysis, and Visualisation:**

Sharif Shaaban ^73^ and Andrew R Hesketh ^101^

**Project administration, Samples and logistics, and Sequencing and analysis:**

Kenneth G Laing ^41^, Irene M Monahan ^41^ and Judith Heaney ^95, 96, 34^

**Project administration, Samples and logistics, and Visualisation:**

Emanuela Pelosi ^97^, Siona Silviera ^97^ and Eleri Wilson-Davies ^97^

**Samples and logistics, Software and analysis tools, and Visualisation:**

Helen Fryer ^5^

**Sequencing and analysis, Software and analysis tools, and Visualization:**

Helen Adams ^4^, Louis du Plessis ^23^, Rob Johnson ^39^, William T Harvey ^53, 42^, Joseph Hughes ^53^, Richard J Orton ^53^, Lewis G Spurgin ^59^, Yann Bourgeois ^81^, Chris Ruis ^102^, Áine O’Toole ^104^, Marina Gourtovaia ^116^ and Theo Sanderson ^116^

**Funding acquisition, and Leadership and supervision:**

Christophe Fraser ^5^, Jonathan Edgeworth ^12^, Judith Breuer ^96, 29^, Stephen L Michell ^105^ and John A Todd ^115^

**Funding acquisition, and Project administration:**

Michaela John ^10^ and David Buck ^115^

**Leadership and supervision, and Metadata curation:**

Kavitha Gajee ^37^ and Gemma L Kay ^75^

**Leadership and supervision, and Project administration:**

David Heyburn ^74^

**Leadership and supervision, and Samples and logistics:**

Themoula Charalampous ^12, 46^, Adela Alcolea-Medina ^32, 112^, Katie Kitchman ^37^, Alan McNally ^43, 93^, David T

Pritchard ^50^, Samir Dervisevic ^58^, Peter Muir ^70^, Esther Robinson ^70, 35^, Barry B Vipond ^70^, Newara A Ramadan ^78^,

Christopher Jeanes ^90^, Danni Weldon ^116^, Jana Catalan ^118^ and Neil Jones ^118^

**Leadership and supervision, and Sequencing and analysis:**

Ana da Silva Filipe ^53^, Chris Williams ^74^, Marc Fuchs ^77^, Julia Miskelly ^77^, Aaron R Jeffries ^105^, Karen Oliver ^116^ and Naomi R Park ^116^

**Metadata curation, and Samples and logistics:**

Amy Ash ^1^, Cherian Koshy ^1^, Magdalena Barrow ^7^, Sarah L Buchan ^7^, Anna Mantzouratou ^7^, Gemma Clark ^15^, Christopher W Holmes ^16^, Sharon Campbell ^17^, Thomas Davis ^21^, Ngee Keong Tan ^22^, Julianne R Brown ^29^, Kathryn A Harris ^29, 2^, Stephen P Kidd ^33^, Paul R Grant ^34^, Li Xu-McCrae ^35^, Alison Cox ^38, 63^, Pinglawathee Madona ^38, 63^, Marcus Pond ^38, 63^, Paul A Randell ^38, 63^, Karen T Withell ^48^, Cheryl Williams ^51^, Clive Graham ^60^, Rebecca Denton-Smith ^62^, Emma Swindells ^62^, Robyn Turnbull ^62^, Tim J Sloan ^67^, Andrew Bosworth ^70, 35^, Stephanie Hutchings ^70^, Hannah M Pymont ^70^, Anna Casey ^76^, Liz Ratcliffe ^76^, Christopher R Jones ^79, 105^, Bridget A Knight ^79, 105^, Tanzina Haque ^80^, Jennifer Hart ^80^, Dianne Irish-Tavares ^80^, Eric Witele ^80^, Craig Mower ^86^, Louisa K Watson ^86^, Jennifer Collins ^89^, Gary Eltringham ^89^, Dorian Crudgington ^98^, Ben Macklin ^98^, Miren Iturriza-Gomara ^107^, Anita O Lucaci ^107^ and Patrick C McClure ^113^

**Metadata curation, and Sequencing and analysis:**

Matthew Carlile ^18^, Nadine Holmes ^18^, Christopher Moore ^18^, Nathaniel Storey ^29^, Stefan Rooke ^73^, Gonzalo Yebra ^73^, Noel Craine ^74^, Malorie Perry ^74^, Nabil-Fareed Alikhan ^75^, Stephen Bridgett ^77^, Kate F Cook ^84^, Christopher Fearn ^84^, Salman Goudarzi ^84^, Ronan A Lyons ^88^, Thomas Williams ^104^, Sam T Haldenby ^107^, Jillian Durham ^116^ and Steven Leonard ^116^

**Metadata curation, and Software and analysis tools:**

Robert M Davies ^116^

**Project administration, and Samples and logistics:**

Rahul Batra ^12^, Beth Blane ^20^, Moira J Spyer ^30, 95, 96^, Perminder Smith ^32, 112^, Mehmet Yavus ^85, 109^, Rachel J Williams ^96^, Adhyana IK Mahanama ^97^, Buddhini Samaraweera ^97^, Sophia T Girgis ^102^, Samantha E Hansford ^109^, Angie Green ^115^, Charlotte Beaver ^116^, Katherine L Bellis ^116, 102^, Matthew J Dorman ^116^, Sally Kay ^116^, Liam Prestwood ^116^ and Shavanthi Rajatileka ^116^

**Project administration, and Sequencing and analysis:**

Joshua Quick ^43^

**Project administration, and Software and analysis tools:**

Radoslaw Poplawski ^43^

**Samples and logistics, and Sequencing and analysis:**

Nicola Reynolds ^8^, Andrew Mack ^11^, Arthur Morriss ^11^, Thomas Whalley ^11^, Bindi Patel ^12^, Iliana Georgana ^24^, Myra Hosmillo ^24^, Malte L Pinckert ^24^, Joanne Stockton ^43^, John H Henderson ^65^, Amy Hollis ^65^, William Stanley ^65^, Wen C Yew ^65^, Richard Myers ^72^, Alicia Thornton ^72^, Alexander Adams ^74^, Tara Annett ^74^, Hibo Asad ^74^, Alec Birchley ^74^, Jason Coombes ^74^, Johnathan M Evans ^74^, Laia Fina ^74^, Bree Gatica-Wilcox ^74^, Lauren Gilbert ^74^, Lee Graham ^74^, Jessica Hey ^74^, Ember Hilvers ^74^, Sophie Jones ^74^, Hannah Jones ^74^, Sara Kumziene-Summerhayes ^74^, Caoimhe McKerr ^74^, Jessica Powell ^74^, Georgia Pugh ^74^, Sarah Taylor ^74^, Alexander J Trotter ^75^, Charlotte A Williams ^96^, Leanne M Kermack ^102^, Benjamin H Foulkes ^109^, Marta Gallis ^109^, Hailey R Hornsby ^109^, Stavroula F Louka ^109^, Manoj Pohare ^109^, Paige Wolverson ^109^, Peijun Zhang ^109^, George MacIntyre-Cockett ^115^, Amy Trebes ^115^, Robin J Moll ^116^, Lynne Ferguson ^117^, Emily J Goldstein ^117^, Alasdair Maclean ^117^ and Rachael Tomb ^117^

**Samples and logistics, and Software and analysis tools:**

Igor Starinskij ^53^

**Sequencing and analysis, and Software and analysis tools:**

Laura Thomson ^5^, Joel Southgate ^11, 74^, Moritz UG Kraemer ^23^, Jayna Raghwani ^23^, Alex E Zarebski ^23^, Olivia Boyd ^39^, Lily Geidelberg ^39^, Chris J Illingworth ^52^, Chris Jackson ^52^, David Pascall ^52^, Sreenu Vattipally ^53^, Timothy M Freeman ^109^, Sharon N Hsu ^109^, Benjamin B Lindsey ^109^, Keith James ^116^, Kevin Lewis ^116^, Gerry Tonkin-Hill ^116^ and Jaime M Tovar-Corona ^116^

**Sequencing and analysis, and Visualisation:**

MacGregor Cox ^20^

**Software and analysis tools, and Visualisation:**

Khalil Abudahab ^14, 116^, Mirko Menegazzo ^14^, Ben EW Taylor MEng ^14, 116^, Corin A Yeats ^14^, Afrida Mukaddas ^53^, Derek W Wright ^53^, Leonardo de Oliveira Martins ^75^, Rachel Colquhoun ^104^, Verity Hill ^104^, Ben Jackson ^104^, JT McCrone ^104^, Nathan Medd ^104^, Emily Scher ^104^ and Jon-Paul Keatley ^116^

**Leadership and supervision:**

Tanya Curran ^3^, Sian Morgan ^10^, Patrick Maxwell ^20^, Ken Smith ^20^, Sahar Eldirdiri ^21^, Anita Kenyon ^21^, Alison H Holmes ^38, 57^, James R Price ^38, 57^, Tim Wyatt ^69^, Alison E Mather ^75^, Timofey Skvortsov ^77^ and John A Hartley ^96^

**Metadata curation:**

Martyn Guest ^11^, Christine Kitchen ^11^, Ian Merrick ^11^, Robert Munn ^11^, Beatrice Bertolusso ^33^, Jessica Lynch ^33^, Gabrielle Vernet ^33^, Stuart Kirk ^34^, Elizabeth Wastnedge ^56^, Rachael Stanley ^58^, Giles Idle ^64^, Declan T Bradley ^69, 77^, Nicholas F Killough ^69^, Jennifer Poyner ^79^ and Matilde Mori ^110^

**Project administration:**

Owen Jones ^11^, Victoria Wright ^18^, Ellena Brooks ^20^, Carol M Churcher ^20^, Laia Delgado Callico ^20^, Mireille Fragakis ^20^, Katerina Galai ^20, 70^, Andrew Jermy ^20^, Sarah Judges ^20^, Anna Markov ^20^, Georgina M McManus ^20^, Kim S Smith ^20^, Peter M D Thomas-McEwen ^20^, Elaine Westwick ^20^, Stephen W Attwood ^23^, Frances Bolt ^38, 57^, Alisha Davies ^74^, Elen De Lacy ^74^, Fatima Downing ^74^, Sue Edwards ^74^, Lizzie Meadows ^75^, Sarah Jeremiah ^97^, Nikki Smith ^109^ and Luke Foulser ^116^

**Samples and logistics:**

Amita Patel ^12^, Louise Berry ^15^, Tim Boswell ^15^, Vicki M Fleming ^15^, Hannah C Howson-Wells ^15^, Amelia Joseph ^15^, Manjinder Khakh ^15^, Michelle M Lister ^15^, Paul W Bird ^16^, Karlie Fallon ^16^, Thomas Helmer ^16^, Claire L McMurray ^16^, Mina Odedra ^16^, Jessica Shaw ^16^, Julian W Tang ^16^, Nicholas J Willford ^16^, Victoria Blakey ^17^, Veena Raviprakash ^17^, Nicola Sheriff ^17^, Lesley-Anne Williams ^17^, Theresa Feltwell ^20^, Luke Bedford ^26^, James S Cargill ^27^, Warwick Hughes ^27^, Jonathan Moore ^28^, Susanne Stonehouse ^28^, Laura Atkinson ^29^, Jack CD Lee ^29^, Dr Divya Shah ^29^, Natasha Ohemeng-Kumi ^32, 112^, John Ramble ^32, 112^, Jasveen Sehmi ^32, 112^, Rebecca Williams ^33^, Wendy Chatterton ^34^, Monika Pusok ^34^, William Everson ^37^, Anibolina Castigador ^44^, Emily Macnaughton ^44^, Kate El Bouzidi ^45^, Temi Lampejo ^45^, Malur Sudhanva ^45^, Cassie Breen ^47^, Graciela Sluga ^48^, Shazaad SY Ahmad ^49, 70^, Ryan P George ^49^, Nicholas W Machin ^49, 70^, Debbie Binns ^50^, Victoria James ^50^, Rachel Blacow ^55^, Lindsay Coupland ^58^, Louise Smith ^59^, Edward Barton ^60^, Debra Padgett ^60^, Garren Scott ^60^, Aidan Cross ^61^, Mariyam Mirfenderesky ^61^, Jane Greenaway ^62^, Kevin Cole ^64^, Phillip Clarke ^67^, Nichola Duckworth ^67^, Sarah Walsh ^67^, Kelly Bicknell ^68^, Robert Impey ^68^, Sarah Wyllie ^68^, Richard Hopes ^70^, Chloe Bishop ^72^, Vicki Chalker ^72^, Ian Harrison ^72^, Laura Gifford ^74^, Zoltan Molnar ^77^, Cressida Auckland ^79^, Cariad Evans ^85, 109^, Kate Johnson ^85, 109^, David G Partridge ^85, 109^, Mohammad Raza ^85, 109^, Paul Baker ^86^, Stephen Bonner ^86^, Sarah Essex ^86^, Leanne J Murray ^86^, Andrew I Lawton ^87^, Shirelle Burton-Fanning ^89^, Brendan AI Payne ^89^, Sheila Waugh ^89^, Andrea N Gomes ^91^, Maimuna Kimuli ^91^, Darren R Murray ^91^, Paula Ashfield ^92^, Donald Dobie ^92^, Fiona Ashford ^93^, Angus Best ^93^, Liam Crawford ^93^, Nicola Cumley ^93^, Megan Mayhew ^93^, Oliver Megram ^93^, Jeremy Mirza ^93^, Emma Moles-Garcia ^93^, Benita Percival ^93^, Megan Driscoll ^96^, Leah Ensell ^96^, Helen L Lowe ^96^, Laurentiu Maftei ^96^, Matteo Mondani ^96^, Nicola J Chaloner ^99^, Benjamin J Cogger ^99^, Lisa J Easton ^99^, Hannah Huckson ^99^, Jonathan Lewis ^99^, Sarah Lowdon ^99^, Cassandra S Malone ^99^, Florence Munemo ^99^, Manasa Mutingwende ^99^, Roberto Nicodemi ^99^, Olga Podplomyk ^99^, Thomas Somassa ^99^, Andrew Beggs ^100^, Alex Richter ^100^, Claire Cormie ^102^, Joana Dias ^102^, Sally Forrest ^102^, Ellen E Higginson ^102^, Mailis Maes ^102^, Jamie Young ^102^, Rose K Davidson ^103^, Kathryn A Jackson ^107^, Alexander J Keeley ^109^, Jonathan Ball ^113^, Timothy Byaruhanga ^113^, Joseph G Chappell ^113^, Jayasree Dey ^113^, Jack D Hill ^113^, Emily J Park ^113^, Arezou Fanaie ^114^, Rachel A Hilson ^114^, Geraldine Yaze ^114^ and Stephanie Lo ^116^

**Sequencing and analysis:**

Safiah Afifi ^10^, Robert Beer ^10^, Joshua Maksimovic ^10^, Kathryn McCluggage ^10^, Karla Spellman ^10^, Catherine Bresner ^11^, William Fuller ^11^, Angela Marchbank ^11^, Trudy Workman ^11^, Ekaterina Shelest ^13, 81^, Johnny Debebe ^18^, Fei Sang ^18^, Sarah Francois ^23^, Bernardo Gutierrez ^23^, Tetyana I Vasylyeva ^23^, Flavia Flaviani ^31^, Manon Ragonnet-Cronin ^39^, Katherine L Smollett ^42^, Alice Broos ^53^, Daniel Mair ^53^, Jenna Nichols ^53^, Kyriaki Nomikou ^53^, Lily Tong ^53^, Ioulia Tsatsani ^53^, Sarah O’Brien ^54^, Steven Rushton ^54^, Roy Sanderson ^54^, Jon Perkins ^55^, Seb Cotton ^56^, Abbie Gallagher ^56^, Elias Allara ^70, 102^, Clare Pearson ^70, 102^, David Bibby ^72^, Gavin Dabrera ^72^, Nicholas Ellaby ^72^, Eileen Gallagher ^72^, Jonathan Hubb ^72^, Angie Lackenby ^72^, David Lee ^72^, Nikos Manesis ^72^, Tamyo Mbisa ^72^, Steven Platt ^72^, Katherine A Twohig ^72^, Mari Morgan ^74^, Alp Aydin ^75^, David J Baker ^75^, Ebenezer Foster-Nyarko ^75^, Sophie J Prosolek ^75^, Steven Rudder ^75^, Chris Baxter ^77^, Sílvia F Carvalho ^77^, Deborah Lavin ^77^, Arun Mariappan ^77^, Clara Radulescu ^77^, Aditi Singh ^77^, Miao Tang ^77^, Helen Morcrette ^79^, Nadua Bayzid ^96^, Marius Cotic ^96^, Carlos E Balcazar ^104^, Michael D Gallagher ^104^, Daniel Maloney ^104^, Thomas D Stanton ^104^, Kathleen A Williamson ^104^, Robin Manley ^105^, Michelle L Michelsen ^105^, Christine M Sambles ^105^, David J Studholme ^105^, Joanna Warwick-Dugdale ^105^, Richard Eccles ^107^, Matthew Gemmell ^107^, Richard Gregory ^107^, Margaret Hughes ^107^, Charlotte Nelson ^107^, Lucille Rainbow ^107^, Edith E Vamos ^107^, Hermione J Webster ^107^, Mark Whitehead ^107^, Claudia Wierzbicki ^107^, Adrienn Angyal ^109^, Luke R Green ^109^, Max Whiteley ^109^, Emma Betteridge ^116^, Iraad F Bronner ^116^, Ben W Farr ^116^, Scott Goodwin ^116^, Stefanie V Lensing ^116^, Shane A McCarthy ^116, 102^, Michael A Quail ^116^, Diana Rajan ^116^, Nicholas M Redshaw ^116^, Carol Scott ^116^, Lesley Shirley ^116^ and Scott AJ Thurston ^116^

**Software and analysis tools:**

Will Rowe ^43^, Amy Gaskin ^74^, Thanh Le-Viet ^75^, James Bonfield ^116^, Jennifier Liddle ^116^ and Andrew Whitwham ^116^

**1** Barking, Havering and Redbridge University Hospitals NHS Trust, **2** Barts Health NHS Trust, **3** Belfast Health & Social Care Trust, **4** Betsi Cadwaladr University Health Board, **5** Big Data Institute, Nuffield Department of Medicine, University of Oxford, **6** Blackpool Teaching Hospitals NHS Foundation Trust, **7** Bournemouth University, **8** Cambridge Stem Cell Institute, University of Cambridge, **9** Cambridge University Hospitals NHS Foundation Trust, **10** Cardiff and Vale University Health Board, **11** Cardiff University, **12** Centre for Clinical Infection and Diagnostics Research, Department of Infectious Diseases, Guy’s and St Thomas’ NHS Foundation Trust, **13** Centre for Enzyme Innovation, University of Portsmouth, **14** Centre for Genomic Pathogen Surveillance, University of Oxford, **15** Clinical Microbiology Department, Queens Medical Centre, Nottingham University Hospitals NHS Trust, **16** Clinical Microbiology, University Hospitals of Leicester NHS Trust, **17** County Durham and Darlington NHS Foundation Trust, **18** Deep Seq, School of Life Sciences, Queens Medical Centre, University of Nottingham, **19** Department of Infectious Diseases and Microbiology, Cambridge University Hospitals NHS Foundation Trust, **20** Department of Medicine, University of Cambridge, **21** Department of Microbiology, Kettering General Hospital, **22** Department of Microbiology, South West London Pathology, **23** Department of Zoology, University of Oxford, **24** Division of Virology, Department of Pathology, University of Cambridge, **25** East Kent Hospitals University NHS Foundation Trust, **26** East Suffolk and North Essex NHS Foundation Trust, **27** East Sussex Healthcare NHS Trust, **28** Gateshead Health NHS Foundation Trust, **29** Great Ormond Street Hospital for Children NHS Foundation Trust, **30** Great Ormond Street Institute of Child Health (GOS ICH), University College London (UCL), **31** Guy’s and St. Thomas’ Biomedical Research Centre, **32** Guy’s and St. Thomas’ NHS Foundation Trust, **33** Hampshire Hospitals NHS Foundation Trust, **34** Health Services Laboratories, **35** Heartlands Hospital, Birmingham, **36** Hub for Biotechnology in the Built Environment, Northumbria University, **37** Hull University Teaching Hospitals NHS Trust, **38** Imperial College Healthcare NHS Trust, **39** Imperial College London, **40** Infection Care Group, St George’s University Hospitals NHS Foundation Trust, **41** Institute for Infection and Immunity, St George’s University of London, **42** Institute of Biodiversity, Animal Health & Comparative Medicine, **43** Institute of Microbiology and Infection, University of Birmingham, **44** Isle of Wight NHS Trust, **45** King’s College Hospital NHS Foundation Trust, **46** King’s College London, **47** Liverpool Clinical Laboratories, **48** Maidstone and Tunbridge Wells NHS Trust, **49** Manchester University NHS Foundation Trust, **50** Microbiology Department, Buckinghamshire Healthcare NHS Trust, **51** Microbiology, Royal Oldham Hospital, **52** MRC Biostatistics Unit, University of Cambridge, **53** MRC-University of Glasgow Centre for Virus Research, **54** Newcastle University, **55** NHS Greater Glasgow and Clyde, **56** NHS Lothian, **57** NIHR Health Protection Research Unit in HCAI and AMR, Imperial College London, **58** Norfolk and Norwich University Hospitals NHS Foundation Trust, **59** Norfolk County Council, **60** North Cumbria Integrated Care NHS Foundation Trust, **61** North Middlesex University Hospital NHS Trust, **62** North Tees and Hartlepool NHS Foundation Trust, **63** North West London Pathology, **64** Northumbria Healthcare NHS Foundation Trust, **65** Northumbria University, **66** NU-OMICS, Northumbria University, **67** Path Links, Northern Lincolnshire and Goole NHS Foundation Trust, **68** Portsmouth Hospitals University NHS Trust, **69** Public Health Agency, Northern Ireland, **70** Public Health England, **71** Public Health England, Cambridge, **72** Public Health England, Colindale, **73** Public Health Scotland, **74** Public Health Wales, **75** Quadram Institute Bioscience, **76** Queen Elizabeth Hospital, Birmingham, **77** Queen’s University Belfast, **78** Royal Brompton and Harefield Hospitals, **79** Royal Devon and Exeter NHS Foundation Trust, **80** Royal Free London NHS Foundation Trust, **81** School of Biological Sciences, University of Portsmouth, **82** School of Health Sciences, University of Southampton, **83** School of Medicine, University of Southampton, **84** School of Pharmacy & Biomedical Sciences, University of Portsmouth, **85** Sheffield Teaching Hospitals NHS Foundation Trust, **86** South Tees Hospitals NHS Foundation Trust, **87** Southwest Pathology Services, **88** Swansea University, **89** The Newcastle upon Tyne Hospitals NHS Foundation Trust, **90** The Queen Elizabeth Hospital King’s Lynn NHS Foundation Trust, **91** The Royal Marsden NHS Foundation Trust, **92** The Royal Wolverhampton NHS Trust, **93** Turnkey Laboratory, University of Birmingham, **94** University College London Division of Infection and Immunity, **95** University College London Hospital Advanced Pathogen Diagnostics Unit, **96** University College London Hospitals NHS Foundation Trust, **97** University Hospital Southampton NHS Foundation Trust, **98** University Hospitals Dorset NHS Foundation Trust, **99** University Hospitals Sussex NHS Foundation Trust, **100** University of Birmingham, **101** University of Brighton, **102** University of Cambridge, **103** University of East Anglia, **104** University of Edinburgh, **105** University of Exeter, **106** University of Kent, **107** University of Liverpool, **108** University of Oxford, **109** University of Sheffield, **110** University of Southampton, **111** University of St Andrews, **112** Viapath, Guy’s and St Thomas’ NHS Foundation Trust, and King’s College Hospital NHS Foundation Trust, **113** Virology, School of Life Sciences, Queens Medical Centre, University of Nottingham, **114** Watford General Hospital, **115** Wellcome Centre for Human Genetics, Nuffield Department of Medicine, University of Oxford, **116** Wellcome Sanger Institute, **117** West of Scotland Specialist Virology Centre, NHS Greater Glasgow and Clyde, **118** Whittington Health NHS Trust

